# A diverse high-fibre plant-based dietary intervention improves gut microbiome composition, gut symptoms, energy and hunger in healthy adults: a randomised controlled trial

**DOI:** 10.1101/2024.07.02.24309816

**Authors:** Alice C. Creedon, Hannah Bernard, Federica Amati, Nicola Segata, Meg Wallace, Alberto Arrè, Harry A. Smith, Alex Platts, William J. Bulsiewicz, Kate M. Bermingham, Joan Capdevila Pujol, Elisa Piperni, Ana Roomans Ledo, Claire Johnson, Catherine Caro, Nafisa Karimjee, Inbar Linenberg, Francesca Giordano, Richard Davies, Jonathan Wolf, Francesco Asnicar, Tim D. Spector, Sarah E. Berry

**Affiliations:** Zoe Ltd, London, UK; Department of Nutritional Sciences, King’s College London, London, UK; Department of Twins Research and Genetic Epidemiology, King’s College London, London, UK; Department of Cellular, Computational and Integrative Biology (CIBIO), University of Trento, Trento, Italy; IEO, Istituto Europeo di Oncologia IRCSS, Milan, Italy; School of Medicine, Emory University, Atlanta, GA 30322, USA; School of Population Health, Royal College of Surgeons in Ireland, Dublin, Ireland

## Abstract

Diets low in diverse fibre-rich plant foods are a major factor in the rise of chronic diseases globally. The BIOME study (NCT06231706) was a 6-week, parallel design randomised controlled trial in 399 healthy adults in the UK, investigating a simple dietary intervention containing 30+ whole-food ingredients high in plant polyphenolic compounds, fibre and micronutrients. Participants were randomised to the primary intervention (prebiotic blend; 30g/d) or control (bread croutons; 28g/d; isocaloric functional equivalent) or a daily probiotic (*L. rhamnosu*s). The primary outcome was change in ‘favourable’ and ‘unfavourable’ microbiome species compared to control, secondary outcomes included changes in blood metabolites, gut symptoms, stool output, anthropometric measures, subjective hunger, sleep, energy and mood. A crossover test meal challenge sub-study was conducted in 34 participants, investigating postprandial glucose responses, subjective hunger, satiety and mood.

In the 349 male and female participants (mean age 50yrs) included in the analysis (intention-to-treat), self-reported adherence was high (> 98% for all treatments). Following the prebiotic blend, significant improvements were seen in the change and ranking of ‘favourable’ and ‘unfavourable’ species as well as beta diversity (weighted-UniFrac measure), but not in the control or probiotic group. There were significantly greater improvements in self reported indigestion, constipation, heartburn, flatulence and energy, following the prebiotic vs control, and hunger following the prebiotic vs probiotic. Addition of the prebiotic to a high carbohydrate test meal challenge resulted in significant improvements in subjective hunger, fullness, and energy (3h incremental area under the curve). No other significant differences between groups were observed.

This prebiotic blend is a simple dietary strategy that benefits gut microbiome composition, gut symptoms and self-reported energy and hunger.

**Graphical abstract:** 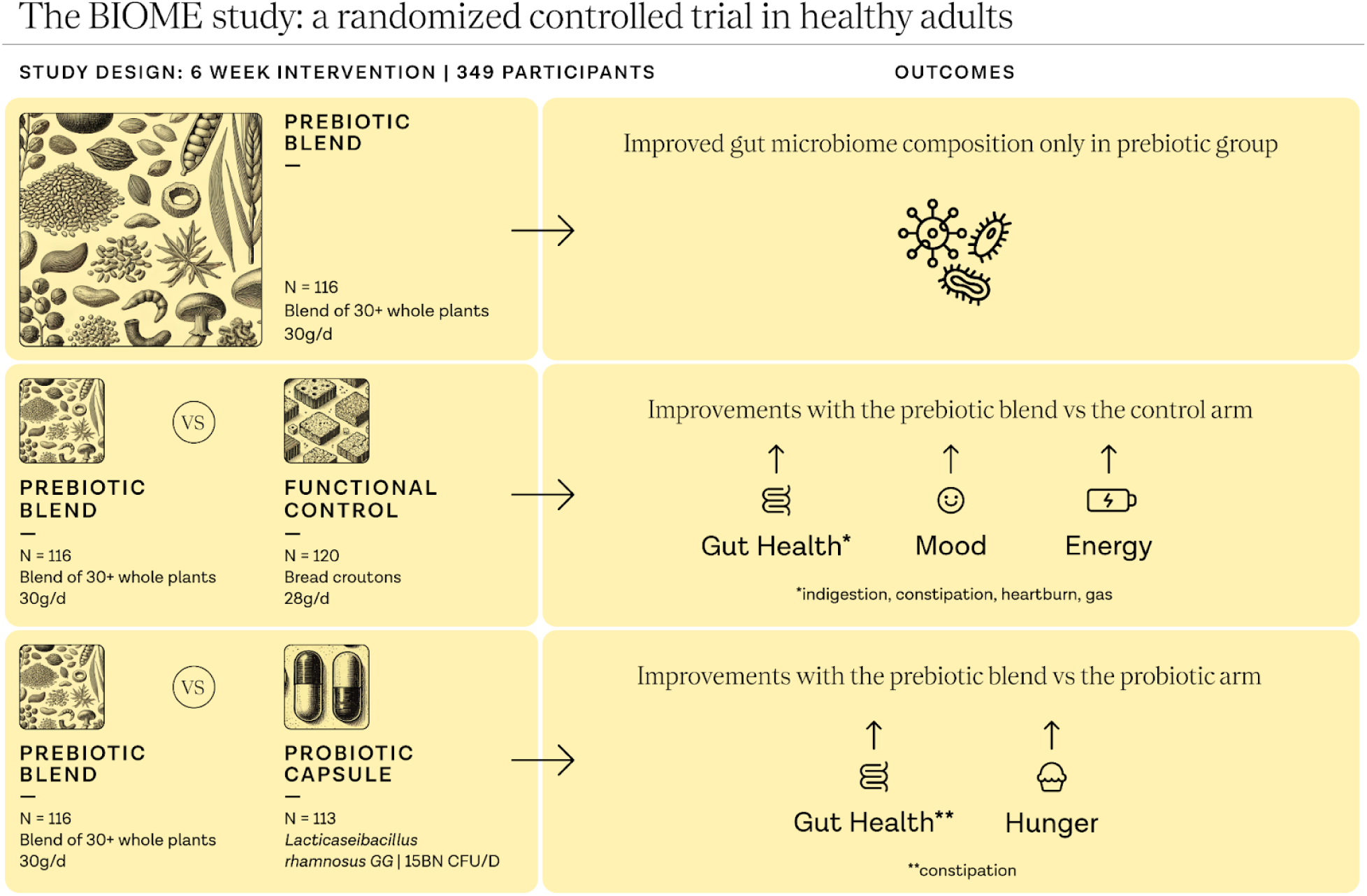

## Main

Despite global public health guidelines, diet remains a key modifiable risk factor for mortality and morbidity from chronic diseases including obesity, type 2 diabetes, cardiovascular disease and cancer ^1, 2^. Public health recommendations to increase intake of plant-based foods that have documented benefits for human health^3, 4^, remain unmet^5^. Additionally, individuals consuming western diets do not consume enough fibre, with 91% in the UK^5^ and 92% in the US^6^ failing to meet national recommended intakes. Further, there is a lack of guidance around increased diversity of plant-based food consumption for the benefit of health, despite strong mechanistic rationale and promising evidence from observational studies^7–9^. However, modifying dietary behaviour is challenging, particularly as a result of modern lifestyles in which people typically have less time to prepare fresh foods^10^, lack nutritional education^11^ and therefore there is a greater demand for convenient choices that are typically less healthy^12^. There is an urgent need for simple dietary strategies aimed at improving health in the current food environment.

Global public health guidelines and research efforts to date have mainly focused on the health impact of the quantity of fruits, vegetables and wholegrains^2, 13^. Increasingly, dietary patterns shown to improve health outcomes are reported to do so in part by modulating intestinal microbial communities^14, 15^. These diets are rich in prebiotic compounds (fermentable fibre, polyphenols) and foods containing probiotic bacteria (fermented foods) with both primarily derived from plant-based food ingredients. While prebiotics and probiotics can be administered in supplement form, their introduction into the diet in foods in which they are naturally present has the combined benefit of provision of additional plant-based micronutrients, proteins, and bio-actives (e.g. polyphenols and other phytochemicals) while preserving the food matrix and micronutrient synergies that exist in whole foods^16^.

The relationship between diet, the gut microbiome and human health is well-documented in observational cohort studies^17, 18^. Dietary intake influences both the composition and diversity of the human gut microbial communities^19^, and in turn these communities and their metabolites contribute towards physiological processes involved in both health and disease states^20–23^. Extensive research in this area has led to the establishment of the gut bacteria as a key mediator in the impact of diet on health and disease processes^24^. Plant-based diets and increased intakes of plant food groups, such as wholegrains, have been shown to modulate gut microbiome composition resulting in positive health outcomes in both healthy adults^25^ and those at increased risk of adverse cardiometabolic health^26, 27^. While causal links between the gut microbiome and diet-related disease continue to be investigated, it is clear that dietary interventions targeting the microbiome have potential to influence the development and treatment of diet-related disease, and the maintenance of health throughout the life course^28^.

Advances in our knowledge and understanding of the gut microbiome has highlighted plant-based diversity as an additional mechanism behind the biological impact of plants on human health^18, 29^. Edible plants and the health benefits they provide extend beyond fruits and vegetables to include legumes, wholegrains, nuts and seeds, herbs and spices. Consuming a diverse range of plant foods has a strong mechanistic rationale for improving health, including greater distribution of micronutrient intake and exposure to a greater range of diverse fibres and polyphenols, that impact gastrointestinal and cardiometabolic health in different ways^30^. A growing body of observational evidence indicates a link between diversity of plant food consumption and the gut microbial community^18, 31^, and the mechanisms by which diet-related changes in gut microbiota diversity and composition contribute to the metabolic health of the host, and chronic cardiometabolic disease processes^32, 33^.

While a growing number of food products are enriched with fibre, or additional nutrients targeting gut microbiome composition and health, few do so using a wide diversity of whole plant foods. Therefore, to address the lack of simple fibre-rich and plant diverse dietary strategies, that are minimally processed and target gut microbiome and associated health outcomes, we designed a prebiotic blend combining more than thirty whole plant ingredients chosen based on their content of diverse fibres and prebiotic compounds (including fruits/vegetables (n=6), mushrooms (n=8), herbs (n=3), nuts (n=3), seeds (n=6), spices (n=2), wholegrains (n=2)), and providing a range of micronutrients, unsaturated fats and polyphenols.

The BIOME (Biotics Influence on Microbiome Ecosystem) study investigated the effect of this prebiotic blend on both chronic and postprandial health outcomes. In a 6-week parallel design randomised controlled trial (**Figure 1a**) we tested the hypothesis that the prebiotic blend will improve gut microbiome composition (primary) and metabolic health (secondary) in comparison to a functional control. To further investigate the efficacy of the intervention on microbiome and health outcomes, we included an active control arm (single-strain probiotic containing *Lacticaseibacillus rhamnosus GG*) to test the hypothesis that the prebiotic blend will improve microbiome composition and metabolic health in comparison to a probiotic supplement. Finally, we conducted a sub-study in a subset of participants, to elucidate the postprandial health effects of the prebiotic blend on glycaemia, and subjective hunger and satiety responses (**Figure 1b**).

**Figure 1.**
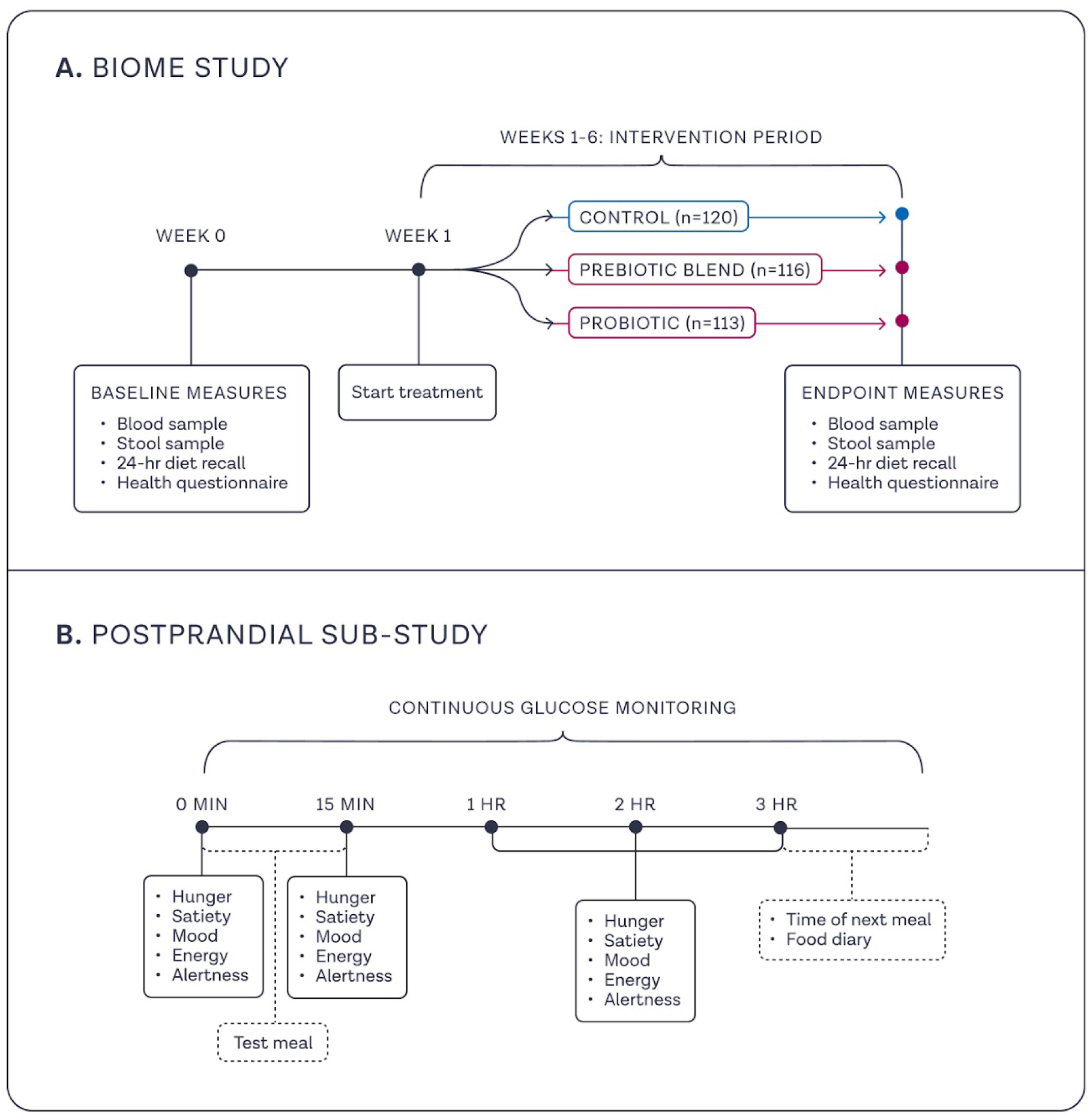
BIOME study design. **A** In the BIOME study, participants were randomly assigned to consume a prebiotic blend (n = 116; 30g/d), a single-strain probiotic capsule (*L.rhamnosus* 15 billion CFU) (n = 113; 1 capsule/d) or bread croutons (n = 120; 28g/d, isocaloric functional control) for 6 weeks. Study outcomes were assessed using health questionnaires, 24-hr dietary recalls, blood and stool samples collected at baseline and 6-weeks. **B** In the postprandial sub-study, a subset of participants who completed the control arm of the BIOME study were invited to take part in a randomised crossover design postprandial test meal challenge (n=34) in which they consumed a breakfast consisting of white bread and low fat spread (57g carbohydrate) with or without the prebiotic blend (30g) in duplicate, separated by a 2-day washout period. Study outcomes were assessed using continuous glucose monitors (CGMs), visual analogue scales and food records completed on each test day.

## Results

### Study participant characteristics

Between 26th February and 22nd April 2024, 8,017 volunteers were screened for initial eligibility, of which 399 participants were randomly assigned to the primary intervention (prebiotic blend; n = 133), active control (probiotic capsule; n = 133) or functional control (bread croutons; n = 133) groups. Fifty participants did not meet the second eligibility screening following randomisation, resulting in 349 participants included in the intention-to-treat analysis set; summarised in the Consolidated Standards of Reporting Trials (CONSORT) diagram (**Figure 2**). In the acute postprandial sub-study, 39 participants who completed the control arm of the chronic phase, and opted to take part were randomly assigned to one of six meal orders. A total of 34 participants completed the postprandial sub-study and were included in the analysis.

**Figure 2.**
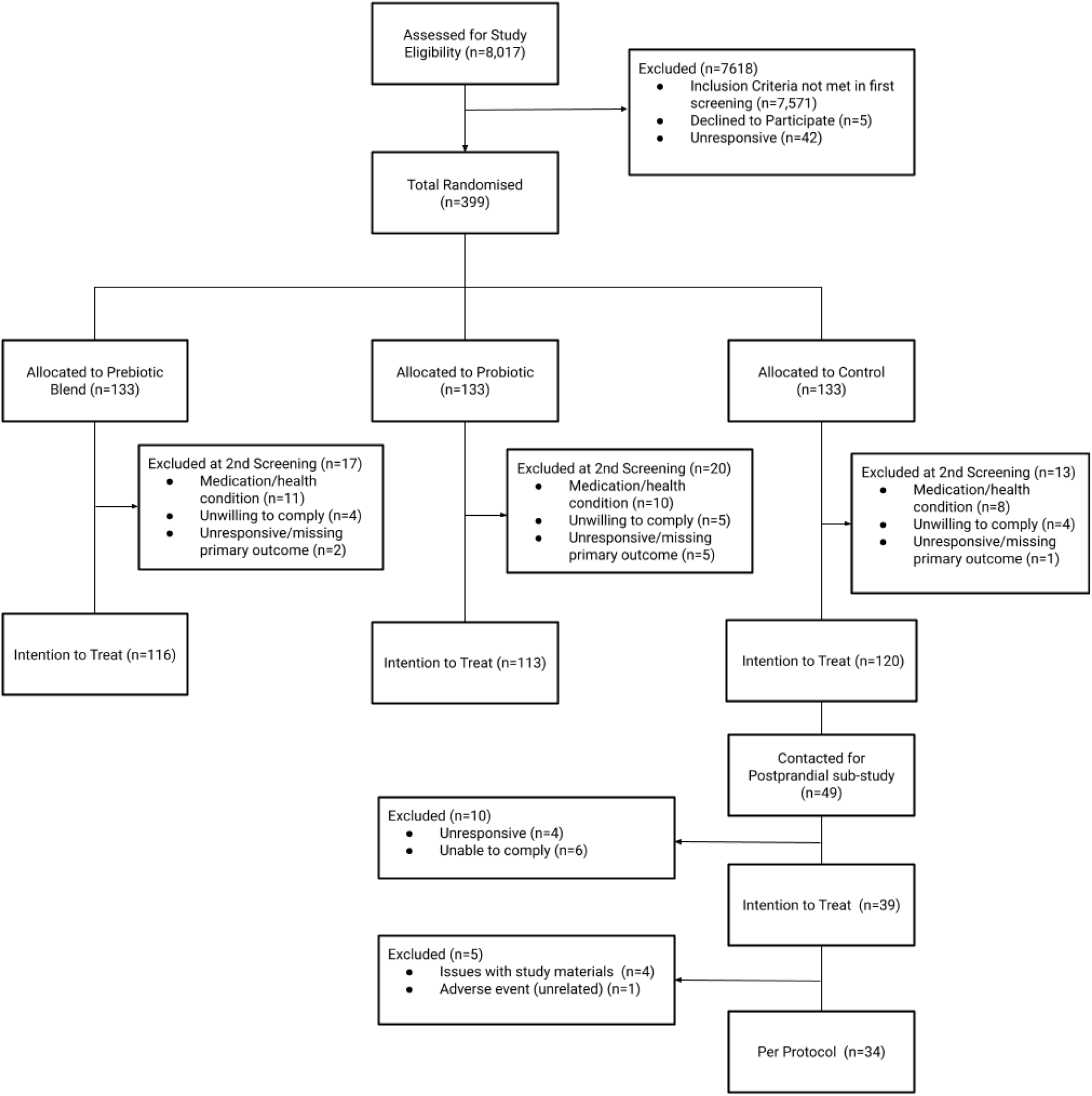
CONSORT Diagram. CONSORT, Consolidated Standards of Reporting Trials

At baseline, there were no significant differences in participant characteristics between groups (**Table 1**). Participants had a median age of 51.4 years (interquartile range, IQR 12.35), a median body mass index (BMI) of 25.9 kg/m^2^ (IQR 5.9) and 75% were female. Diet quality was within population ranges (mean Healthy Eating Index (HEI) 69.0, s.d. 8.6), and participants reported 135 minutes (IQR 180) of moderate-vigorous physical activity per week. Participants fell within the inclusion criteria for daily intake of fibre (median 16.4 g/d (IQR 5.3)) and fermented foods (median 0.3 servings (IQR 0.4)). There were no significant differences between groups (prebiotic vs control; prebiotic vs probiotic) for age, sex physical activity or HEI at baseline (*p* > 0.05 for all).

**Table 1.**
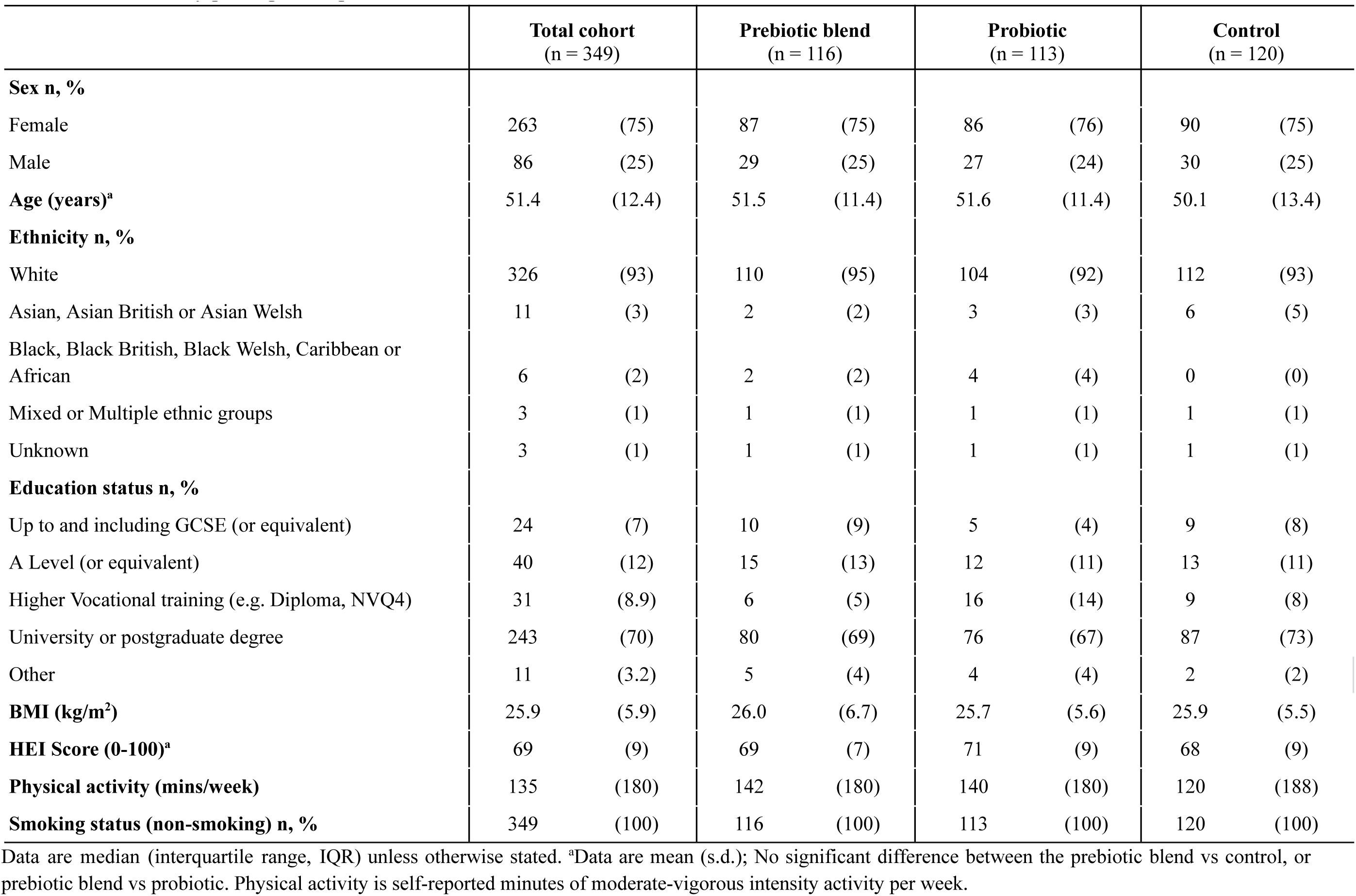
BIOME study participant disposition.

|  | **Total cohort**<br>(n = 349) |  | **Prebiotic blend**<br>(n = 116) |  | **Probiotic**<br>(n = 113) |  | **Control**<br>(n = 120) |  |
| --- | --- | --- | --- | --- | --- | --- | --- | --- |
| **Sex n, %** |  |  |  |  |  |  |  |  |
| Female | 263 | (75) | 87 | (75) | 86 | (76) | 90 | (75) |
| Male | 86 | (25) | 29 | (25) | 27 | (24) | 30 | (25) |
| **Age (years)<sup>a</sup>** | 51.4 | (12.4) | 51.5 | (11.4) | 51.6 | (11.4) | 50.1 | (13.4) |
| **Ethnicity n, %** |  |  |  |  |  |  |  |  |
| White | 326 | (93) | 110 | (95) | 104 | (92) | 112 | (93) |
| Asian, Asian British or Asian Welsh | 11 | (3) | 2 | (2) | 3 | (3) | 6 | (5) |
| Black, Black British, Black Welsh, Caribbean or African | 6 | (2) | 2 | (2) | 4 | (4) | 0 | (0) |
| Mixed or Multiple ethnic groups | 3 | (1) | 1 | (1) | 1 | (1) | 1 | (1) |
| Unknown | 3 | (1) | 1 | (1) | 1 | (1) | 1 | (1) |
| **Education status n, %** |  |  |  |  |  |  |  |  |
| Up to and including GCSE (or equivalent) | 24 | (7) | 10 | (9) | 5 | (4) | 9 | (8) |
| A Level (or equivalent) | 40 | (12) | 15 | (13) | 12 | (11) | 13 | (11) |
| Higher Vocational training (e.g. Diploma, NVQ4) | 31 | (8.9) | 6 | (5) | 16 | (14) | 9 | (8) |
| University or postgraduate degree | 243 | (70) | 80 | (69) | 76 | (67) | 87 | (73) |
| Other | 11 | (3.2) | 5 | (4) | 4 | (4) | 2 | (2) |
| **BMI (kg/m<sup>2</sup>)** | 25.9 | (5.9) | 26.0 | (6.7) | 25.7 | (5.6) | 25.9 | (5.5) |
| **HEI Score (0-100)<sup>a</sup>** | 69 | (9) | 69 | (7) | 71 | (9) | 68 | (9) |
| **Physical activity (mins/week)** | 135 | (180) | 142 | (180) | 140 | (180) | 120 | (188) |
| **Smoking status (non-smoking) n, %** | 349 | (100) | 116 | (100) | 113 | (100) | 120 | (100) |

Data are median (interquartile range, IQR) unless otherwise stated. <sup>a</sup>Data are mean (s.d.); No significant difference between the prebiotic blend vs control, or prebiotic blend vs probiotic. Physical activity is self-reported minutes of moderate-vigorous intensity activity per week.

### Adherence to treatment and habitual diet

Self-reported adherence to assigned treatments in the intention to treat (ITT) cohort (n = 341; n = 8 missing data from all weekly questionnaires) was high overall (98.8%) and across groups; prebiotic blend (98.1%), probiotic (99.2%), control (99.2%). Participants were instructed to maintain a habitual background diet, monitored by completion of 24 hr dietary recalls at baseline and 6-weeks. Participants in the prebiotic blend group had marginally greater energy intake from total sugar (mean ± s.d.; 16% ± 7%) in comparison to the probiotic group (15% ± 5%; *p* = 0.038, ANCOVA). There were no other differences in energy or macronutrient intake between groups (**Supplementary Table 1**).

### Microbiome analysis

The pre-specified primary outcome of the study was improvement in microbiome composition measured using the ranking of prevalent microbiome species associated with cardiometabolic health. This ranking was developed by our group^34^, to identify and prioritise microbial species most likely affecting host health either in a positive or negative way in over 34,000 individuals^34^. We identified the prevalent species (classified using species level genome bins) at baseline and 6-weeks post interventions and then tested if the relative abundance values between the two time points were significantly different (paired Wilcoxon signed-rank test; FDR adjusted *p*-values < 0.01). In the ITT cohort we identified *n* = 57 species that were significantly different over time in the prebiotic blend group; *n* = 4 species in the probiotic group; and *n* = 14 species in the control group. Of these significant species, we then distinguished them according to whether they increased or decreased their relative abundance at 6-weeks, and tested whether the significantly increasing (or decreasing) species had significantly different “ZOE Microbiome Ranking 2024 (Cardiometabolic Health)”. Rank values closer to 0 are indicative of ‘favourable’ species associated with better health predictions^34^, while ranks closer to 1 indicate ‘unfavourable’ species associated with poorer health outcomes. In the prebiotic blend group, the median rank of decreasing species (0.659) was significantly higher (‘unfavourable’) than the median rank of increasing species (0.408, ‘favourable’), indicating that the prebiotic supplementation impacts the microbiome by increasing species associated with favourable cardiometabolic health markers while decreasing those associated with less favourable cardiometabolic health markers (*p* = 0.007; Mann-Whitney U-test; **Figure 3a,c**). This significant effect was not seen in either the probiotic group (median rank (decreasing) = 0.604; median rank (increasing) = 0.694; *p* = 0.500) or the control group (median rank (decreasing) = 0.604; median rank (increasing) = 0.519; *p* = 0.555) (**Figure 3a**). As well as the distributions changing in the prebiotic arm, the significantly changing species had different prevalence patterns over time. Increasing (generally ‘favourable’) species maintained or increased their prevalence (from 74.1% to 76.9% median prevalence, Wilcoxon paired test *p* = 0.001), while decreasing generally ‘unfavourable’ species became undetectable in many individuals (from 61.3% to 44.3% median prevalence, Wilcoxon paired test *p* < 0.001) (**Figure 3d**).

**Figure 3.**
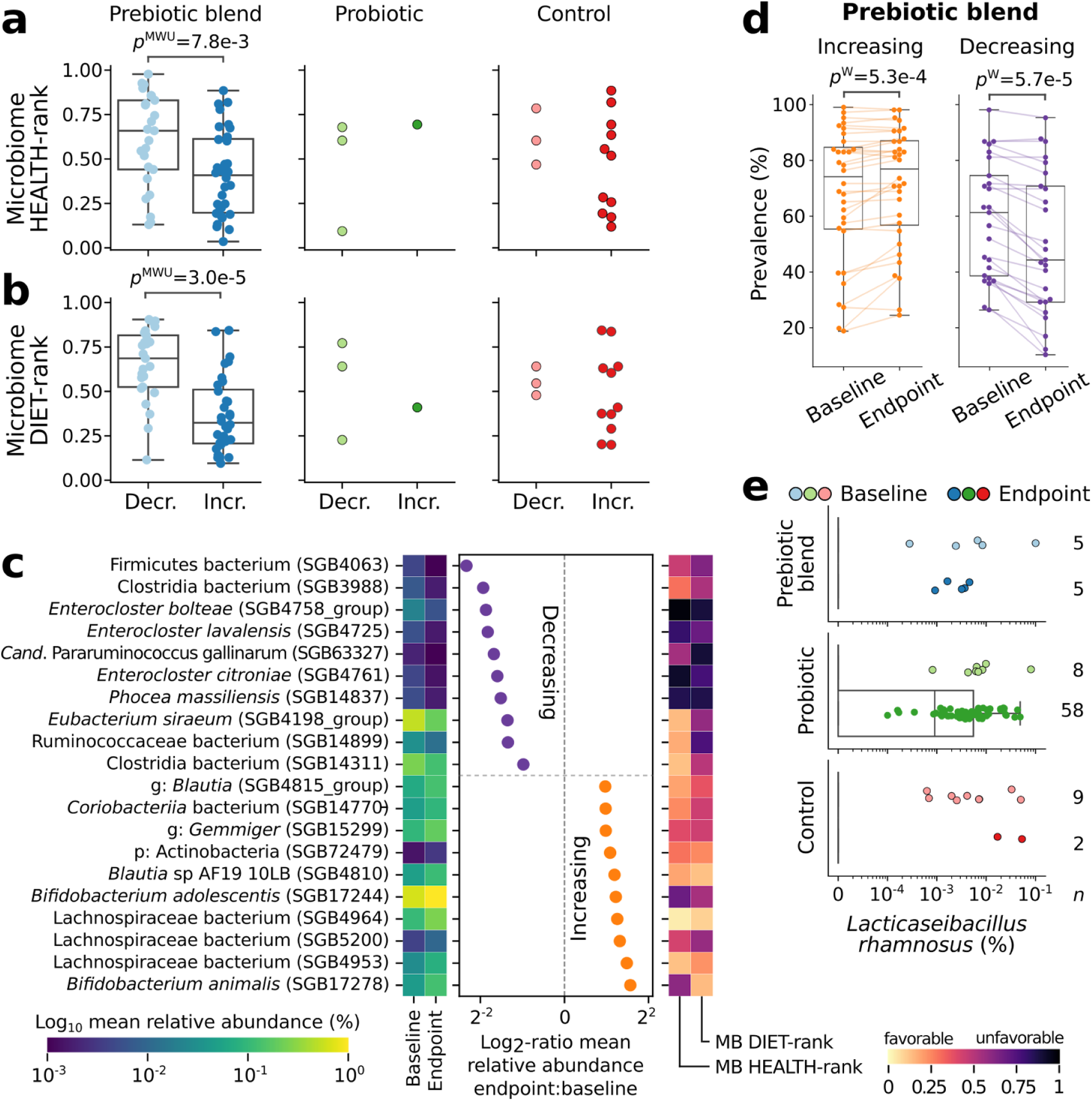
Effect of the prebiotic blend on the gut microbiome. **a,b** In each group, species with a significant change in their relative abundance at baseline compared to endpoint were identified and categorised into those that decrease or increase at endpoint. For these species, we evaluated their ZOE MB Health and Diet ranks. Only the prebiotic blend group showed statistically significant differences in the ZOE MB ranks between decreasing and increasing species. This shows that species that decrease at endpoint have higher ZOE MB Health and Diet ranks, meaning these are more ‘unfavourable’ species. Species that increase in relative abundance have lower ranks meaning these are more ‘favourable species’. **c** The top 10 species that increase or decrease in the prebiotic blend group, according to their log_2_-fold change at endpoint are reported, showing their relative abundance values at both time points (heatmap to the left) and their change in prevalence. The ZOE MB Health and Diet ranks are reported (heatmap to the right). **d** For the significantly increasing (left) and decreasing (right) species of the prebiotic blend group, we reported their prevalence at baseline and endpoint, showing that not only these species increase or decrease their relative abundance values, but the ‘favourable’ species that increase tend also to be more prevalent, i.e., more present across individuals (from 74.1% to 76.9% median prevalence). While the ‘unfavourable’ species that decrease in abundance also tend to be much less prevalent among individuals (from 61.3% to 44.3% median prevalence). **e** We verified the presence and relative abundance of the probiotic species *Lacticaseibacillus rhamnosus* (formerly known as *Lactobacillus rhamnosus)* among the three groups and only the probiotic group showed a significantly larger number of individuals from which we were able to identify *L. rhamnosus* in their gut microbiome at 6-weeks.

Similarly, to explore the effect of the interventions on microbiome species associated with markers of diet quality, we evaluated the identified significant species according to their “ZOE Microbiome Ranking 2024 (Diet)” ^34^. In the prebiotic blend group, the median rank of decreasing species (0.686, ‘unfavourable’) was significantly higher than the median rank of increasing species (0.323, ‘favourable’), indicating that increasing species were those associated with favourable diet quality indices while decreasing species were those associated with less favourable diet quality indices (*p* < 0.001; Mann-Whitney U-test; **Figure 3b**). This significant effect was not seen in either the probiotic group (median rank (decreasing) = 0.641; median rank (increasing) = 0.410; *p* = 1.0) or the control group (median rank (decreasing) = 0.546; median rank (increasing) = 0.410; *p* = 0.55) (**Figure 3b**).

As traditional gut microbiome analysis, we also calculated the weighted-UniFrac measure of beta-diversity and both species richness and Shannon alpha diversity measures **(Supplementary Figure 1).** For the weighted-UniFrac measure, there was no separation between the baseline microbiome composition across the three groups (PERMANOVA *p*-value = 0.584), while endpoint microbiome compositions showed significant differences (PERMANOVA *p*-value = 0.020). PERMANOVA analysis performed within each group comparing baseline with endpoint microbiome composition, showed significant differences only for the prebiotic blend group (prebiotic blend *p*-value = 0.030 probiotic *p*-value = 0.327, control *p*-value = 0.059). For alpha diversity, we could detect only slight changes across all the groups, Shannon’s diversity index tended to increase at endpoint but this was only significant in the probiotic group, (Wilcoxon’s *p*-value = 0.020), while richness decreased significantly in the prebiotic blend group only (Wilcoxon’s *p*-value = 0.001).

Finally as a measure of adherence we investigated the presence of the probiotic *L. rhamnosus* across the groups at baseline and 6-weeks. As expected, only the probiotic group showed a significantly larger number of individuals from which we were able to identify *L. rhamnosus* in their gut microbiome at 6-weeks compared to baseline (from 5 to 58 participants) (**Figure 3f**).

### Secondary outcomes

#### Subjective measures

Hunger, energy and mood were assessed using visual analogue scales administered online (0-100; digital VAS) and gastrointestinal symptoms were assessed using the Gastrointestinal Symptom Rating Scale (GSRS)^35^ at baseline and 6-weeks. For the primary comparison (prebiotic vs control) a greater proportion of participants reported improvements in energy (50.5% vs 37.3%); happiness (44.6% vs 30%); severity of indigestion domain symptoms (55.1% vs 36.4%); severity of constipation domain symptoms (34.6%, vs 24.5%), severity of flatulence (37.4% vs 23.6%); severity of heartburn (17.8% vs 11.8%); and total gastrointestinal symptoms (69.2% vs 56.4%) following the prebiotic vs the control. Estimates of effect size are reported in **Figure 4**. Stool frequency was significantly different following the prebiotic blend in comparison to control at 6-weeks, with a smaller proportion of the prebiotic blend group reporting stool frequencies of “Three or four times per week” (3.5%), vs control (14.2%) (**Supplementary Table 2**). There were no differences between the groups in self-reported sleep quality (**Supplementary Table 2**). However, when assessed as the proportion of participants who improved sleep quality, there was a significantly greater proportion who improved sleep quality following the prebiotic blend vs the control (34.9% vs 20%; chi-square *p* = 0.014). Anthropometric (self reported) measures (waist circumference and weight), stool consistency, acne, sleep quantity, and remaining subjective emotions and gastrointestinal symptoms did not differ significantly between groups (**Table 2; Supplementary Table 2**).

**Figure 4.**
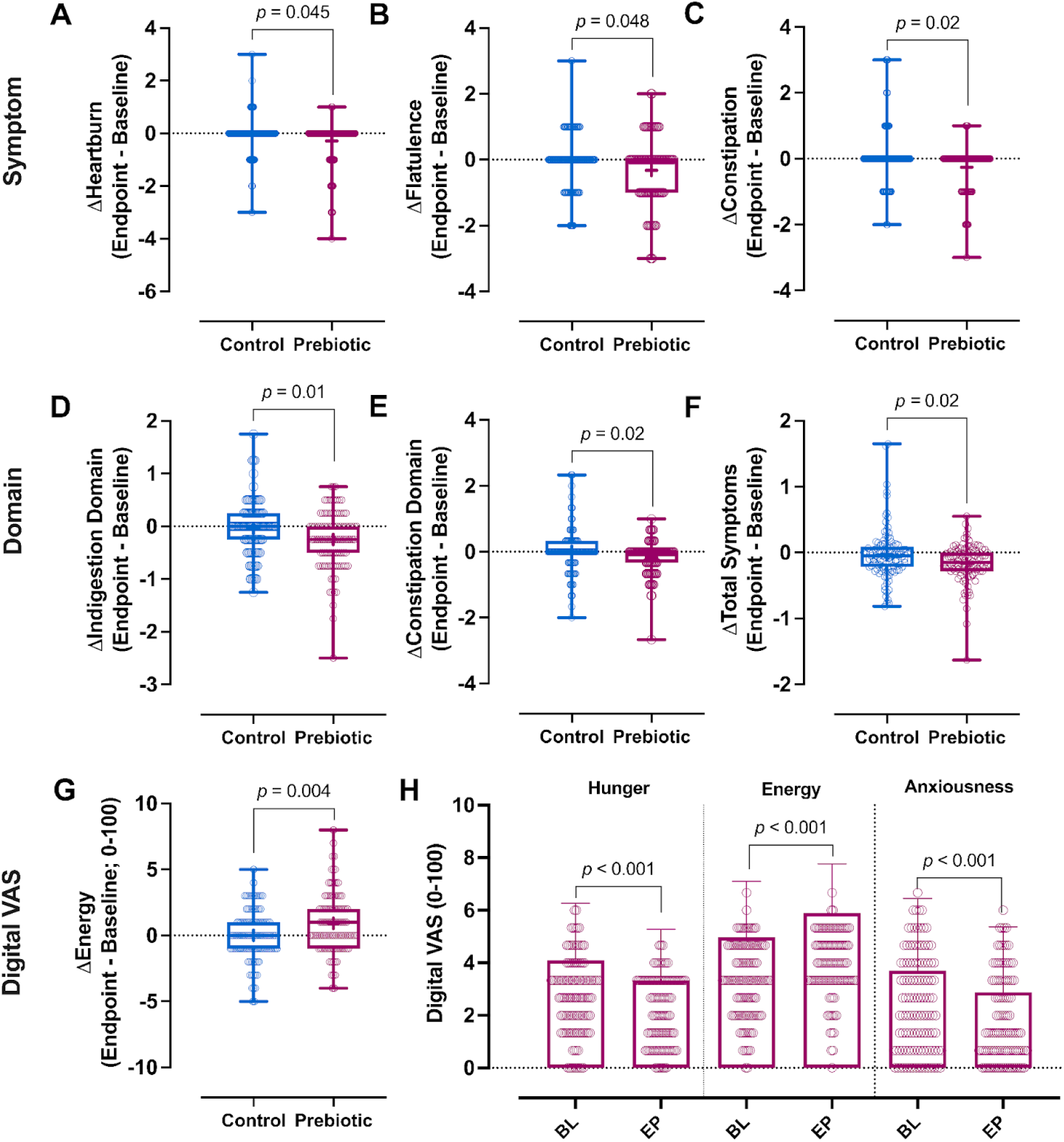
Changes in selected secondary outcomes in the BIOME study. **a-c** Changes in severity of individual gastrointestinal symptoms including heartburn (**a**), flatulence (**b**) and constipation (**c**) in the prebiotic blend (n = 107) and control (n = 110) groups. **d-e** Changes in severity of gastrointestinal symptom domains including indigestion (**d**) and constipation (**e**) in the prebiotic blend (n = 107) and control (n = 110) groups. **f** Change in severity of total gastrointestinal symptoms in the prebiotic blend (n = 107) and control (n = 110) groups. **g** Changes in subjective energy ratings (digital visual analogue scale, digital VAS; 0-100) from baseline to 6-weeks in the prebiotic blend (n = 107) and control (n = 110) groups. **h** Subjective ratings of hunger, energy and anxiousness in the prebiotic blend group (n = 107) at baseline and endpoint. Data presented are median, first and third quartile (box plots) and range (error bars) for all. The mean value is also presented (+). *P*-values are the result of Mann-Whitney U-tests (a-g) or Wilcoxon signed-rank test (h). Prebiotic blend (pink), and control (blue) for all.

**Table 2.** Selected secondary outcomes in the BIOME study.

|  | Prebiotic blend |  |  | Probiotic |  |  | Control |  |  | *p-values* |  |
| --- | --- | --- | --- | --- | --- | --- | --- | --- | --- | --- | --- |
|  | *Baseline*<br>(n = 116) | *Endpoint*<br>(n = 107) | *Δ baseline-end* | *Baseline*<br>(n = 113) | *Endpoint*<br>(n = 107) | *Δ baseline-end* | *Baseline*<br>(n = 120) | *Endpoint*<br>(n = 110) | *Δ baseline-end* | Prebiotic<br>blend<br>vs control | Prebiotic<br>blend<br>vs probiotic |
| **Subjective feelings**<br>**(Digital VAS)** |  |  |  |  |  |  |  |  |  |  |  |
| Energy | 4.97 ± 2.12 | 5.90 ± 1.87 | 0.93 (0.57, 1.28) | 5.09 ± 1.96 | 5.91 ± 1.71 | 0.85 (0.56, 1.14) | 5.16 ± 2.11 | 5.26 ± 1.97 | 0.01 (-0.29, 0.30) | **0.004** | 0.990 |
| Hunger | 4.09 ± 2.18 | 3.33 ± 1.95 | -0.86 (-1.24, -0.48) | 3.74 ± 2.31 | 3.68 ± 2.23 | 0.04 (-0.30, 0.37) | 3.96 ± 2.22 | 3.47 ± 2.15 | -0.44 (-0.81, -0.06) | 0.230 | **0.006** |
| Happiness | 6.30 ± 1.80 | 6.61 ± 1.55 | 0.30 (0.03, 0.57) | 6.11 ± 1.88 | 6.59 ± 1.92 | 0.48 (0.21, 0.75) | 5.94 ± 1.95 | 5.83 ± 1.87 | -0.22 (-0.48, 0.04) | **0.020** | 0.500 |
| Anxious | 3.71 ± 2.74 | 2.88 ± 2.50 | -0.99 (-1.38, -0.61) | 3.63 ± 2.41 | 2.78 ± 2.51 | -0.75 (-1.05, -0.44) | 3.72 ± 2.48 | 3.11 ± 2.65 | -0.55 (-0.93, -0.16) | 0.260 | 0.660 |
| **Gastrointestinal symptom severity** |  |  |  |  |  |  |  |  |  |  |  |
| Acid reflux | 1.37 ± 0.90 | 1.16 ± 0.39 | -0.19 (-0.30, -0.09) | 1.21 ± 0.54 | 1.07 ± 0.23 | -0.08 (-0.14, -0.02) | 1.18 ± 0.47 | 1.18 ± 0.52 | 0.00 (-0.08, 0.08) | 0.103 | 0.334 |
| Abdominal pain | 1.31 ± 0.40 | 1.21 ± 0.38 | -0.09 (-0.15, -0.03) | 1.40 ± 0.56 | 1.16 ± 0.28 | -0.23 (-0.31, -0.16) | 1.40 ± 0.63 | 1.32 ± 0.59 | -0.06 (-0.15, 0.03) | 0.818 | 0.081 |
| Indigestion | 1.68 ± 0.62 | 1.44 ± 0.50 | -0.25 (-0.34, -0.16) | 1.74 ± 0.71 | 1.40 ± 0.45 | -0.33 (-0.42, -0.24) | 1.61 ± 0.71 | 1.54 ± 0.66 | -0.04 (-0.12, 0.04) | **0.011** | 0.334 |
| Diarrhoea | 1.38 ± 0.60 | 1.27 ± 0.51 | -0.12 (-0.21, -0.04) | 1.35 ± 0.79 | 1.17 ± 0.41 | -0.15 (-0.27, -0.03) | 1.36 ± 0.56 | 1.31 ± 0.69 | -0.05 (-0.16, 0.06) | 0.963 | 0.612 |
| Constipation | 1.38 ± 0.52 | 1.23 ± 0.40 | -0.17 (-0.25, -0.09) | 1.39 ± 0.50 | 1.35 ± 0.57 | -0.03 (-0.13, 0.07) | 1.44 ± 0.66 | 1.42 ± 0.72 | 0.01 (-0.09, 0.12) | **0.023** | 0.221 |
| **Total symptoms** | 1.42 ± 0.36 | 1.26 ± 0.29 | -0.16 (-0.21, -0.12) | 1.42 ± 0.41 | 1.23 ± 0.24 | -0.16 (-0.22, -0.11) | 1.40 ± 0.41 | 1.35 ± 0.51 | -0.03 (-0.09, 0.03) | **0.018** | 0.971 |

Data were not normally distributed, but are presented as mean±SD or mean (95%CI) to illustrate precise changes between the groups for each outcome (as median values did not indicate the direction); *p*-values are a result of a Mann-Whitney U-test on median change from baseline values; Digital VAS, visual analogue scales (0-100).

For the secondary comparison (prebiotic vs probiotic), there were greater reductions in severity of constipation (individual symptom, −0.26 vs 0.04; *p* = 0.007) (**Supplementary Table 2)**, and hunger (−0.86 vs −0.44; *p* = 0.006) (**Table 2**) following the prebiotic vs the probiotic. Additionally, there was a significantly greater proportion of participants reporting improved sleep quality following the prebiotic blend vs the probiotic (34.9% vs 19.6%; chi-square *p* = 0.012). There were no other clinically relevant differences between groups (**Table 2; Supplementary Table 2**).

#### Metabolomics

For both the primary comparison of prebiotic blend vs control and the secondary comparison of the prebiotic blend vs probiotic, there were no clinically relevant differences between groups for changes in metabolites after the intervention. (**Supplementary Table 3**).

### Post-hoc analysis

Subgroup analysis of participants within the top tertile concentrations (using baseline values) of key measures of lipids (apolipoproteinB, ApoB; LDL-cholesterol, LDL-C; Triglycerides, TG), and inflammation (glycoprotein acetyls, GlycA) was performed. Within group analysis in this subgroup comparing baseline vs 6 weeks showed small but statistically significant reductions in the prebiotic group for ApoB (−0.06 mmol/L, *p* = 0.003; n = 33), LDL-C (−0.22 mmol/L; *p* = 0.001; n = 32) and GlycA (−0.04 mmol/; *p* = 0.029; n = 29). Similarly, in the probiotic group there were small reductions in GlycA (−0.04 mmol/; *p* = 0.034; n = 34), LDL-C (−0.04 mmol/; *p* = 0.031; n = 30), and TG (−0.2 mmol/; *p* = 0.022; n = 30), but no significant changes in the control group. There were no significant differences between groups observed for changes in these outcome measures. (**Supplementary Table 4**).

Subgroup analysis of gastrointestinal symptoms was also conducted on participants who reported symptoms at baseline with a severity score ≥2 and only for symptoms that were reported by ≥25% of participants at baseline to ensure adequate sample size (n = 10 symptoms). Within-group analysis in this subgroup comparing baseline vs 6 weeks showed small but statistically significant reductions for all symptoms assessed in each group (*p <* 0.05 for all). Between-group analysis showed significant differences between the prebiotic blend and control for improvements in severity of rumbling stomach (mean (95%CI); −1.0 (−1.2, −0.8) vs −0.5 (−0.8, −0.3); *p* = 0.008) and constipation (−1.0 (−1.2, −0.8) vs −0.45 (−0.8, −0.1); *p* = 0.033) (**Supplementary Table 4**).

### The impact of the prebiotic blend on postprandial glucose responses, energy, hunger and satiety

In the postprandial cross-over sub-study (n = 34) comparing a high carbohydrate standardised breakfast with or without the prebiotic blend, there were no significant differences in postprandial glycaemia (**Supplementary Table 5**). However, the addition of the prebiotic blend to the test meal resulted in significantly greater subjective fullness (by 41.5%; Median [IQR]: 481 [275, 699] vs 328 [191, 573] mm x 3h, *p* = 0.001), meal satisfaction (by 21.6%; mean ± s.d.; 243 ± 290 vs 299 ± 316 mm x 3h, *p* = 0.037), and energy (by 43.3% Median [IQR]: 91 [62, 270] vs 192 [119, 328] mm x 3h, *p* = 0.03) and lower hunger (by −16.9%, mean ± s.d.; 56 ± 76 vs 21 ± 33 mm x 3h, *p* = 0.03), desire to eat (by −70.9%, mean ± s.d.; 75 ± 78 vs 25 ± 37 mm x 3h, *p* = 0.003) and prospective consumption (by −54.2%, mean ± s.d.; 48 ± 63 vs 15 ± 24 mm x 3h, *p* = 0.01); **Figure 5**. Energy and macronutrient intake at the next meal (**Supplementary Table 5**) was not significantly different following test meals; with the exception of fiber for which there was a small but significantly lower intake following the high carbohydrate meal with the prebiotic blend, in comparison to the high carbohydrate meal alone (mean ± s.d.; 4.1g ± 3.1 vs 5.1g ± 3.7; *p* = 0.045), potentially as a result of the additional fibre consumed from the test meal with the prebiotic blend (11.9g fibre) vs without the blend (2.9g fibre). Time to the next meal did not differ significantly following test meals (**Supplementary Table 5**). There was significant meal*time interaction for fullness (*p* < 0.05) whereby subjective fullness ratings were higher 60, 120 and 180 mins after consuming the prebiotic blend with the high carbohydrate meal vs by itself (all *p* < 0.05). No other differences in time course analysis between meals for glucose metrics or subjective outcomes were observed (**Supplementary Figures 2-3**).

**Figure 5.**
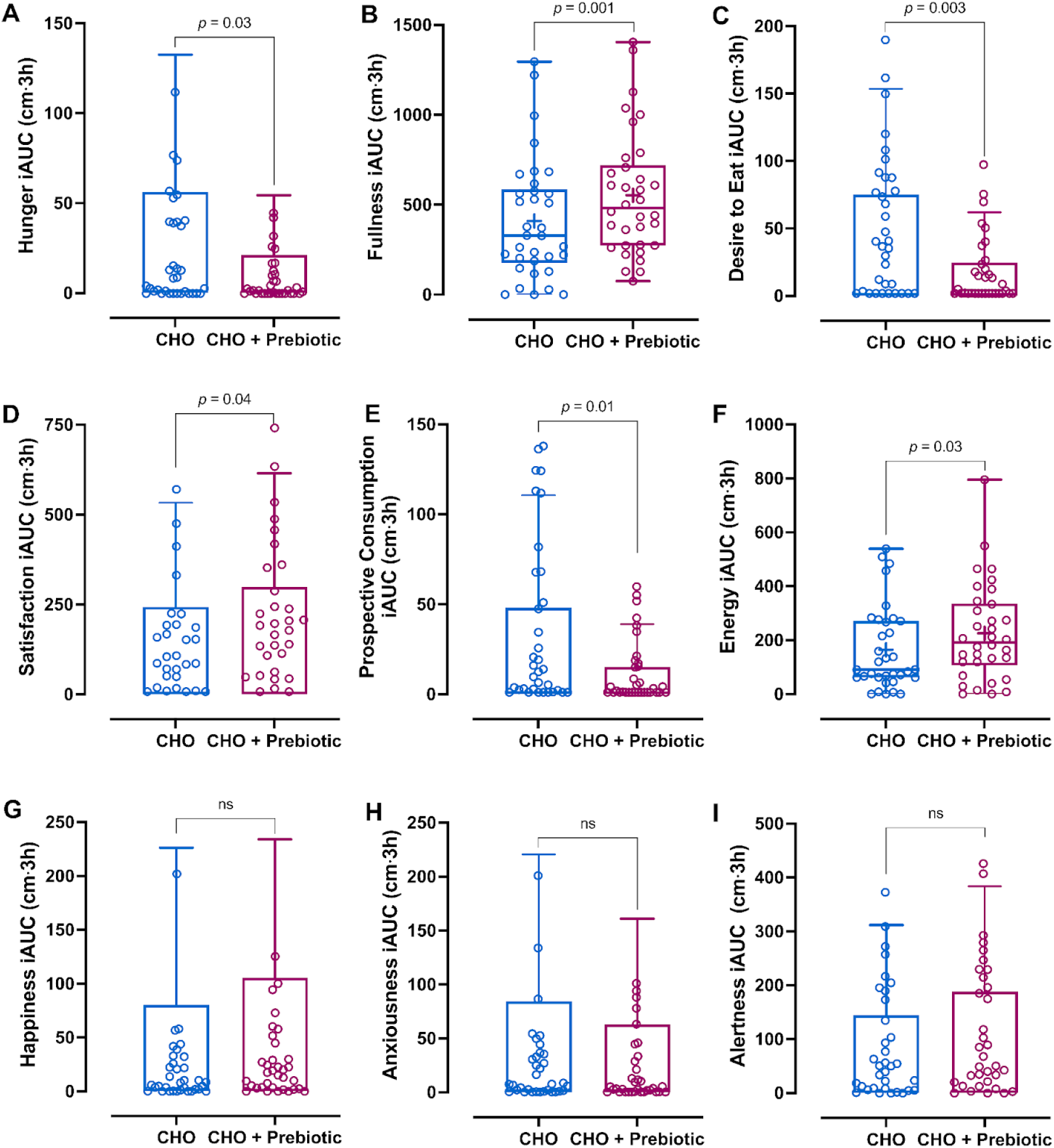
Improvements in postprandial subjective ratings of energy, hunger and satiety following consumption of the prebiotic blend in a postprandial crossover study (n=34) Comparison of subjective ratings of hunger (**a**), fullness (**b**), desire to eat (**c**), satisfaction (**d**), prospective consumption (**e**), energy (**f**), happiness (**g**), anxiousness (**h**), and alertness (**i**) following consumption of a high carbohydrate meal alone (blue) (CHO) or with a prebiotic blend (pink) (CHO + prebiotic) (n = 34, crossover design). Ratings were assessed using visual analogue scales completed at 0hr (immediately before meal consumption, baseline), 15 min, 1hr, 2hr and 3hr. For **a, c, d, e, g-i** data presented are mean 3h incremental area under the curve (iAUC), standard deviation (box plots) and range (error bars). The mean value is also presented (+). *P*-values are the result of a linear mixed-effects model with fixed effects for timepoint, meal, and their interaction, and participant ID as a random intercept. For **b, f** data presented are median 3h incremental area under the curve (iAUC), first and third quartile (box plots) and range (error bars) for all. The mean value is also presented (+). *P*-values are the result of paired Wilcoxon signed-rank tests.

## Discussion

Effective strategies are urgently needed to improve diet quality to reduce the burden of diet-related disease and increase healthy years. In this randomised controlled trial, we demonstrated that a simple, single dietary strategy in the form of a plant-diverse high-fibre prebiotic blend improves gut microbiome composition, gastrointestinal symptoms and subjective feelings of energy, mood and hunger. In addition to the chronic impact of the prebiotic on health over 6 weeks, the postprandial sub-study demonstrated an immediate impact of the prebiotic blend on reductions in hunger and increased satiety and energy.

The increase in energy and reduction in hunger observed in both the chronic and postprandial studies following the prebiotic is pertinent to our current food environment. The food landscape in countries where a western dietary pattern is prevalent, is one of excessive consumption of foods high in sugar, salt and saturated fats, and low in fibre, that typically do not have a high satiating capacity therefore encourage overconsumption^36^. Therefore, while shifts in overall dietary pattern may slowly induce broader health effects, convenient dietary strategies that are low-burden have potential to impact diet quality and therefore diet-related health outcomes in the shorter term and in individuals with demanding lifestyles for whom larger dietary shifts may be unattainable.

Following the prebiotic blend, there were significant changes in the gut microbiome vs both a probiotic and control, with increases in species previously associated with favourable measures of cardiometabolic health and diet and a reduction in species associated with unfavourable measures of health and diet^34^. We also observed a decrease in richness in the prebiotic group. However, with the increased taxonomic resolution available from MetaPhlAn 4.0, previous research has questioned whether traditional diversity metrics are a valid measure of host health^37, 38^. In addition, diversity measures are limited, in that they do not consider whether species present are associated with positive or negative health outcomes.

In accordance with the improvements in gut microbiome species associated with cardiometabolic health^34^ following the prebiotic blend, we observed reductions in measures of inflammation (GlycA) and lipids (ApoB) from baseline to 6-weeks in a sub-group with the highest concentrations of these measures before the intervention. While improvements were small, and of unclear clinical significance in our primarily healthy population, our findings suggest that regular consumption of the blend by individuals at greater cardiometabolic risk may have benefits for metabolic and immune health. Similar results were seen following consumption of the probiotic, indicating the prebiotic blend can exert effects similar to those seen using established interventions targeting the gut microbiome. This is particularly interesting considering previous investigations of the probiotic species used in our study, which have shown beneficial effects on immune health^39^, cognition^40^ and weight loss^41^ in adult participants, outcomes which might therefore warrant investigation in future trials of the health effects of this prebiotic blend. Additionally, the prebiotic resulted in significantly greater improvements in mood and energy compared to the control. This aligns with the growing and promising body of evidence suggesting that diet has the potential to impact brain processes and behaviour via the microbiota gut-brain axis^42^, with RCT’s showing a benefit of prebiotic fibres on mood^43^.

While we did not observe an impact of the prebiotic blend on postprandial glucose concentrations in this population, acute consumption of interventions containing fibre, consumed as a preload, or at the same time as a high carbohydrate meal has been demonstrated to improve postprandial glucose concentrations in individuals with increased cardiometabolic risk^44^ or type 2 diabetes^45, 46^. Thus, it is possible that the additional fibre from prebiotic blend may benefit postprandial gylcaemia in people at greater metabolic risk, an outcome that warrants investigation in future trials. In line with our findings, a meta-analysis of RCTs investigating the impact of dietary prebiotics on postprandial ratings of hunger and satiety reported that prebiotics resulted in greater postprandial satiety^47^.

A strength of this study was the high self-reported adherence to all treatments, in particular indicating that the prebiotic blend was feasible and well tolerated by participants. This study approach also highlights the real world application of the prebiotic blend, in that it can be added to the diet as a “functional swap” for less nutritionally dense products. High adherence indicates that continued use beyond 6 weeks is likely to be acceptable and convenient, and may yield greater shifts in gut microbiome and subsequent benefits for cardiometabolic health.

An added contribution of this research to the field of nutrition was the study design approach. We conducted an exclusively remote RCT, incorporating a 6-week dietary intervention assessing multiple measures of health (gut microbiome, metabolomics and self-reported measures), with both high self-reported adherence and participant retention. We have demonstrated the feasibility of this approach and a potential paradigm shift for dietary intervention trials, in that they can be conducted remotely while maintaining scientific rigour.

Limitations of our study include the absence of a true control food, and therefore lack of blinding of participants to the intervention. This is a common challenge in dietary intervention trials due to the technical limitations of designing a control food that is both nutritionally and functionally similar, but without the active ingredients of the intervention. Therefore we chose a control that was isocaloric and functionally similar, in that the prebiotic blend is designed to be added to meals, in a similar way to croutons. Nonetheless, it is possible that self-reported results may suffer from a degree of subjectivity as a result of participants’ potential awareness of their allocated group (intervention vs control). Additionally, in the postprandial sub-study the interventions were not matched for energy, macronutrients, or carbohydrate load, due to the presence of a small amount of carbohydrate in the prebiotic blend. This design was once again chosen to specifically investigate the impact of the blend on postprandial glycaemia when used as intended, as an addition to a meal. This also considered that the starch profile of the blend was different to that of the bread croutons and therefore the carbohydrate in the blend was not interchangeable with that in the breakfast.

Other limitations include the duration of the intervention (6 weeks) which may have been insufficient to detect clinically meaningful changes in blood metabolites. We also did not include young or elderly individuals, or those suffering from diabetes or severe obesity or many from non-european backgrounds, so we cannot generalise results to all these groups. Although the adherence based on weekly calls was very high, we did not include any objective measure of intake, which is a technical limitation of remote studies, and nutritional studies in a free-living setting.

In conclusion, we present a simple, convenient fibre-rich plant diverse blend of whole food ingredients that can be consumed daily as an addition to the diet, or to replace less nutrient dense alternatives that are designed to add flavour, but are typically nutrient poor. As well as encouraging improvement in diet quality and fibre intake, convenient simple strategies such as the addition of this prebiotic blend provide promising additional benefits to microbiome composition, subjective energy and hunger, and possibly cardiometabolic health.

## Methods

### Study Design

The ZOE BIOME (*Biotics Influence on Microbiome Ecosystem*) Study was a 6-week parallel-designed randomised controlled trial (RCT) conducted exclusively remotely in the UK. In this free-living dietary intervention trial, participants were randomly assigned to receive one of three treatments: (i) a prebiotic blend, consisting of thirty whole-food ingredients high in plant polyphenolic compounds, fibre and micronutrients known to exert prebiotic effects on the gut microbiome (**Supplementary Table 6**); (ii) a single-strain probiotic containing *Lacticaseibacillus rhamnosus GG*, provided in capsule form (active control); or (iii) bread croutons, an energy-matched functional equivalent to the prebiotic blend (functional control) (**Figure 1a**). To test the acute health effects of the prebiotic blend, we conducted a postprandial sub-study in a subgroup of participants. In a crossover design, participants consumed test meals in duplicate in a randomly assigned order. Test meals consisted of a) white bread and low fat spread (control) and b) white bread, low fat spread and prebiotic blend (**Figure 1b**). The study was registered on clincialtrials.gov (NCT06231706) and received ethical approval from King’s College London Research Ethics Committee (HR/DP-23/24-39673). All procedures were carried out in accordance with the Declaration of Helsinki (2013) and Good Clinical Practice.

### Participant selection and randomization

Participants were healthy male and female adults reflective of the average UK population (aged 35-65 y; body mass index (BMI) 18.5 - 40 kg/m^2^; fibre intake <20g/day). Sex was determined using self-reported questionnaires with the following question, ‘Please enter your sex as it was assigned at birth’. Volunteers were excluded from the study if any of the following criteria applied; unable to provide written informed consent through an electronic consent form; unable or unwilling to comply to the study protocol; unwilling to complete study tasks on specified dates; did not complete the Food Frequency Questionnaire (FFQ)^48^ at screening; had previously completed the ZOE Nutrition Product; unwilling to consume study treatments; not based in the UK for the duration of the study; unable to eat the study treatments safely and comfortably (e.g. suffering from inflammatory bowel disease, coeliac disease, Crohn’s disease, irritable bowel syndrome, allergies or intolerances, chronic constipation or chronic diarrhoea); BMI of <18.5 kg/m^2^ or >40 kg/m^2^; following a non-omnivore diet (vegan, vegetarian); high fermented food intake in the previous month (≥ 7 servings per week); fibre intake ≥ 20g/d in the previous month; treatment with medication or products that may impact study outcome measures in the previous 3 months (e.g. antibiotics, non-topical steroids or other immunosuppressive medicines, biologics, probiotics/prebiotics, metformin, chronic use of non-steroidal anti-inflammatory drugs); use of opiate pain medicine for 8 or more days during the previous 3 months; use of proton pump inhibitors for 8 or more days during the previous 3 months; current smoker; suffered from a heart attack, stroke, or major surgery in previous 2 months; received treatment for cancer in the previous 3 months; were pregnant, breastfeeding or planning pregnancy; were suffering from an eating disorder, type 1 or type 2 diabetes mellitus. Participants were recruited to the trial between 12th January and 16th February 2024 by electronic advertisement (emails to the ZOE Health Studies mailing list and ZOE Product Waitlist). Interested volunteers were screened to assess eligibility against the trial inclusion and exclusion criteria in a two part process. First, volunteers who responded to recruitment emails were invited to complete an online screening questionnaire and FFQ. If eligible according to the initial online screening, participants were enrolled in the study and provided electronic informed consent via email with study coordinators. Participants were randomly allocated (ratio 1:1:1) to one of the three treatment groups using a variance minimisation procedure^49^, with sex (male; female), BMI (18.5 - 24.9kg/m^2^; 25 - 40 kg/m^2^), and diet quality (Healthy Eating Index; 0-59; 60-100) as stratification variables. The probability of random assignment (pRand) was set to 0.1^49^. Study coordinators performed randomisation and informed participants of their allocation to treatment via email. The second part of the screening process was conducted as a video welcome call, during which eligibility criteria from the first screening were verified and if participants were identified as not meeting eligibility at this second screening, they were excluded prior to baseline tasks. In addition, study coordinators explained trial procedures. Blinding of participants to the intervention was not possible due to the nature of the test meals (whole foods, for which a placebo that is void of nutrients/properties of interest, but physically similar, is not possible to create). On participant facing materials, we did not indicate which treatment was the intervention of interest, in an attempt to mask participants. Study coordinators were unblinded to the participant’s randomised group, due to the nature of the study (a remote dietary intervention trial). Analysts were blinded to the treatment group for the duration of statistical analysis, by re-coding the treatment groups as groups 1-3.

### Treatments

The nutrient composition of the prebiotic blend and control foods is included in **Supplementary Table 7**. The intervention group received a prebiotic blend (Daily30+; made for ZOE ltd. UK by Indi Supplements, UK; 30g/d) for 6 weeks. The prebiotic blend was provided in generic, unbranded packaging with a label listing major allergens. Participants were instructed to consume the treatment by adding it to meals as part of their usual diet. The active control group received a single-strain probiotic containing *Lacticaseibacillus rhamnosus GG*, provided in capsule form and were instructed to consume 1 capsule daily for 6 weeks. The probiotic was provided in its original packaging with a label listing major allergens covering the front of the package. The functional control group received bread croutons (Tesco Olive Oil and Sea Salt Croutons; Tesco, UK), an energy-matched functional equivalent to the prebiotic blend. The croutons were provided in original packaging with a label listing major allergens covering the ingredient and nutrition information on the back of the packaging. Participants were instructed to consume croutons (28g/d) for 6 weeks by adding them to meals throughout the day.

### Procedures

The study design is summarised in **Figure 1a**. Participants received a study kit via postal delivery containing materials necessary for completing study measurements prior to baseline (week 0) and endpoint (beginning of week 7). All procedures were conducted by participants in their homes.

### Baseline week (week 0)

#### Health Questionnaires

Participants completed health questionnaires administered through an online survey (www.typeform.com; www.surveymonkey.com) for collection of baseline and covariate data including subjective ratings of hunger, energy, and mood, gastrointestinal symptoms, anthropometric measurements (waist circumference, body weight), stool frequency and consistency, sleep (quality and quantity), and physical activity.

#### Dietary intake

To capture habitual dietary intake, participants completed an online 24 hour dietary recall (24hr recall; Intake24^50^) on three specified days during the baseline week. Participants were instructed not to report consumption of their assigned treatment via the 24hr recall to enable assessment of habitual intake only. Adherence to treatment was assessed as described below. The Intake24 tool prompts participants to list all food and drinks consumed the previous day (from midnight to midnight) using free text entry. Foods were then matched to equivalent items using food composition codes in the Intake24 database, the UK Nutrient Databank. Portion size was reported by participants by selection of a single portion size from a range of options accompanied by food photographs within the online questionnaire. Participants were asked to review their entered items and given the option to enter any further intake before submitting their recall.

#### Stool Sample Collection

Stool samples for microbiome analysis were collected by participants at home using the Zymo Research Corporation’s DNA/RNA ShieldTM Fecal Collection Tube containing a buffer (catalogue no. R1101; Zymo Research). The kit contained all the necessary materials for sample collection, along with detailed instructions for use. Participants were instructed to store the sample at room temperature until return by prepaid post to Prebiomics Lab (Trento, Italy).

#### Blood sample collection

Blood samples for metabolomic analysis were collected by participants using the Nightingale Kit® for remote blood collection (Nightingale Health plc, Finland). Participants were instructed to fast overnight before completing the sample collection in line with kit instructions. Upon completion, sample collection devices were stored in return pouches with desiccant and returned via prepaid postal envelope to a receiving laboratory in the UK. The samples were stored at −80 °C upon receipt until shipping to the Nightingale Health laboratory for analysis (Nightingale Health Plc, Helsinki, Finland).

### Participant monitoring and adherence

Participants confirmed completion of primary baseline study tasks via a survey administered at the end of week 0, and again following completion of endpoint tasks. Participants who did not report completion of tasks were contacted via telephone or email. Participants in all three arms were asked to self-report adherence to their allocated treatment by completing a questionnaire administered weekly throughout the study period with the following matrix question, ‘Please fill out the table below to tell us how much of your treatment you consumed each day over the past week’. For the prebiotic blend group, participants were able to select one of the following answer options for each day of the week, ‘0 scoops, 1 scoop, 2 scoops, >2 scoops for each day of the week’ (1 scoop = 15g). For the capsule group, participants were able to select one of the following answer options for each day of the week, ‘0 capsules, 1 capsule, >1 capsule for each day of the week’. For the control group, participants were first asked if they weighed or counted their croutons before being able to select one of the following answer options for each day of the week, ‘0 croutons, 1 crouton, 2 croutons, …22 croutons, > 22 croutons’ or ‘0 grams, 1 gram, 2 grams,…, 28 grams, > 28 grams’. Participants were instructed to maintain their habitual diet during the study; adherence to this instruction was evaluated through 24hr recalls completed at baseline and endpoint.

### Endpoint Measures (week 7)

Endpoint data collection was completed in the 7th week of the study, at which point both groups had consumed their allocated treatments for 6 weeks. All participants completed endpoint measures including health questionnaires, 24hr recall, blood sample and stool sample collection as outlined in the baseline week section above. An additional question was asked at the endpoint only to assess skin improvement.

### Primary outcome measure

The primary outcome of the study was the change in microbiome composition from baseline to the 6-week endpoint, derived from metagenomic analysis of stool samples. Analysis of the primary outcome involved identification of species with a statistically significant difference in terms of relative abundance values from baseline to endpoint, followed by statistical testing of whether the significantly increasing species had significantly higher values of the “ZOE Microbiome Ranking 2024 (Cardiometabolic Health)”^34^ compared to the values of the significantly decreasing species within each group.

### Secondary outcome measures

Secondary outcomes measures were assessed at baseline and following 6-weeks of treatment (i.e. during week 7). Dried blood samples were provided for metabolomic analysis of markers of lipid profile, fatty acids, glucose control and inflammation (full list of metabolites included in the analysis in **Supplementary Table 3)** via high-throughput Nuclear Magnetic Resonance (NMR) metabolomics (Nightingale Health, Helsinki, Finland). Participants were asked to self-report anthropometric measures including body weight (kg), and waist circumference (cm) which was measured by participants using a measuring tape provided in their study kit. Gut symptoms were assessed using the gastrointestinal symptoms rating scale^35^. Frequency of bowel movements was assessed via a single question “On average, how often do you have a bowel movement?” with the following response options: Once a week or less; Twice a week; Three or four times a week; Five or six times a week; Once a day; Twice a day; Three times a day; Four times a day; Five or more times a day. Stool consistency was assessed via the question “Among the seven choices shown in the image, which stool form is the most common/typical that you experience?” and participants responded by indicating their most common stool consistency on the Bristol Stool Form Scale^51^. Subjective feelings (hunger, energy, happiness, anxiety) were assessed via visual analogue scales administered online (digital VAS) with a scale of 0-100^52^. Sleep quality was assessed via the question “During the last 7 days, how would you rate your sleep quality overall?”, adapted from a previously validated question^53^, while sleep quantity data was gathered via the question “During the last 7 days, how many hours of actual sleep did you get at night?”, with response options (hrs): Less than 5; 5-6; 6-7; 7-8; 8-9; 9-10; 10-11; 11-12; More than 12. Skin quality was determined using the question “If you experience acne, has it improved since starting the BIOME study?” with the following response options: Yes; No; Unsure; Not applicable (endpoint only).

### Blood processing and metabolomic analysis

Samples not meeting quality requirements (device not closed, return pouch not closed or sample not sufficient for analysis) were not included in analysis. A total of 106 metabolites were quantified from blood samples; concentrations for 105 biomarkers were quantified as previously described for venous samples^54, 55^. Briefly, approx. 375 mm^2 was taken from the membrane, placed into sodium phosphate buffer (38 mM, pH 7, 10% D2O, 0.04 % sodium 3-(trimethylsilyl)propionate-2,2,3,3-d4 (TSP), and 0.02 % sodium azide), and shaken gently for one hour. For each sample, 520 µL of the extract was transferred into a 5 mm NMR tube for the NMR analysis. For the 106th metabolite (HbA1c) concentration was determined using a Roche cobas c513 analyser with Tina-quant Haemoglobin A1c Third Generation assay. For the analysis, one 6 mm punch was taken from the membrane, placed into a haemolysing reagent (Roche Diagnostics GmbH, Mannheim, Germany), and incubated 30 minutes at room temperature. For each sample, one millilitre of hemolysate was processed in the analyser as per the standard protocol for hemolysate.

### Faecal sampling and microbiome testing

#### DNA extraction and sequencing

DNA was isolated by using the DNeasy 96 PowerSoil Pro QIAcube HT Kit (Qiagen, #47021). The DNA was quantified by using the Quant-iT™ 1X dsDNA Assay Kits, BR (Life Technologies, #Q33267) in combination with the Varioskan LUX Microplate Reader (Thermo Fisher Scientific, #VL0000D0). The DNA was diluted in water for the following library preparation.

#### Library Preparation and Sequencing

The sequencing libraries were prepared with the Illumina DNA Prep, (M) Tagmentation (96 Samples, IPB) kit (Illumina, #20060059) in combination with the Illumina® DNA/RNA UD Indexes Set A, B, C, D, Tagmentation (96 Indexes, 96 Samples) (Cat. #20091654, #20091656, #20091658, #20091660) and the amplified libraries were purified with the double-sided bead purification procedure, as described by the Illumina protocol. Then, libraries concentration (ng/µl) were quantified with the Quant-iT™ 1X dsDNA Assay Kits, HS (Life Technologies, #Q33232) in combination with the Varioskan LUX Microplate Reader (Thermo Fisher Scientific, #VL0000D0). In addition, the base pair length (bp) was evaluated by using the D5000 ScreenTape Assay (Agilent, #5067-5588/9) in combination with the TapeStation 4150 (Agilent Technologies, #G2992AA). By knowing both library concentration and base pair length, it is possible to obtain the correct library volume to pool in the same tube in order to achieve optimal cluster density. The library pool was then quantified with the Qubit 1x dsDNA HS kit (Life Technologies, #Q33231) through the Qubit® 3.0 Fluorometer (Life Technologies, #Q33216) and the base pair length (bp) was evaluated as described before. Finally, the library pools were sequenced using the Novaseq X plus platform (Illumina) at an average depth of 3.75 Gb per sample.

### Metagenome quality control and preprocessing

All sequenced metagenomes were preprocessed using the pipeline implemented in https://github.com/SegataLab/preprocessing. Briefly, the pipeline consists of three steps, the first step involves read-level quality control and removes low-quality reads (Q<20), too short reads (length <75bp), and reads with >2 ambiguous nucleotides. The second step screens for contaminant DNAs using Bowtie 25 with the ‘--sensitive-local’ parameter, allowing confident removal of the phi X 174 Illumina spike-in and human-associated reads (hg19 reference human genome release). The last step consists in splitting and sorting the cleaned reads to create standard forward, reverse and unpaired reads output files for each metagenome (average: 35 ± 13 million reads per sample).

### Microbiome taxonomic profiling

Species-level profiling of the samples was performed with MetaPhlAn 4.0. Default parameters were used for MetaPhlAn, with the following database, “mpa_vJan21_CHOCOPhlAnSGB_202103”. MetaPhlAn 4 taxonomic profiles were used to assess the presence and contribution of the previously identified 50 positively-associated and 50 negatively-associated species with dietary and cardiometabolic health markers^34^. MetaPhlAn 4 taxonomic profiles were analysed to compare microbial compositions among participants and to compute an alpha diversity indices, the number of detected species (“observed richness”) and the number of detected species taking into account their relative abundance (“Shannon’s Diversity Index”). Microbiome taxonomic profiles were also analysed to compare between microbiome samples dissimilarity (beta-diversity) using the unweighted-UniFrac measure.

### Nutrient intake and diet quality

Daily habitual energy and macronutrient intakes were assessed by averaging the energy and macronutrient intakes from three consecutive 24hr dietary recalls at baseline and 6-weeks. Diet quality was assessed by applying the Healthy Eating Index (HEI)^56^.

### Postprandial sub-study

A randomised, controlled, single-blinded 2-phase crossover design study was conducted to determine the effect of the prebiotic blend when consumed alongside a standardised high carbohydrate breakfast (white bread, low fat spread; 60g of available carbohydrate), in comparison to consumption of the high carbohydrate standardised breakfast alone. The sub-study was conducted remotely in the UK. Study outcome measures were postprandial glucose response, subjective ratings of hunger, satiety, mood and energy and amount consumed at next meal. Participants who completed the control arm of the BIOME study were contacted via email and given the option to take part in the postprandial sub-study. Interested participants were sent a participant information sheet detailing the procedures involved in this additional measurement (**Figure 1b**); and completed a welcome video call with study coordinators for explanation of study procedures. Participants provided electronic informed consent prior to enrollment in the sub-study via email with study coordinators. Blinding of participants to the intervention was not possible due to the nature of the test meals (whole foods, for which a placebo that is void of nutrients/properties of interest, but physically similar, is not possible to create). Study coordinators and analysts were blinded to treatment allocation by coding test meals in all data collection documents and databases as “Test meal A” (control) and “Test meal B” (intervention). Treatment sequence was randomised using online software (https://www.sealedenvelope.com/) by an independent researcher. This was performed by randomly assigning participants to one of six possible meal sequences (AABB, ABAB, ABBA, BBAA, BABA, BAAB) using the block randomization service (block sizes of 12)^57^, stratified by biological sex (male, female). Participants were informed of meal sequence via a printed instruction leaflet that was included in their study kit. Study kits were sent to participants via post ahead of the study start date, and contained the following items; continuous glucose monitor (CGM; Abbott Diabetes Care, Alameda CA, USA), food weighing scales (Arc Digital Kitchen Scale; Salter, UK), prebiotic blend (made for ZOE ltd., UK by Indi Supplements, UK), plastic scoop (capacity 15g), study guide, questionnaire booklet, prepaid return envelope, meal sequence leaflet. To ensure fresh food items were within sell-by dates participants were given instructions of exact fresh food items to purchase from local supermarkets; white bread (Warburtons Farmhouse White Bread, Warburtons UK) and low fat spread (Flora Lighter Spread, Flora UK); and received reimbursement for these products. The study took part over a 10-day intervention period. Test meals were consumed in duplicate over 4 test days (days 1, 4, 7 and 10), with each test day separated by a 2-day washout period.

Participants were instructed to apply their CGM on the upper non-dominant arm, the day before their first test day (day 0). An adhesive patch was applied on top of the monitor to ensure secure attachment (Sourceful, Manchester, UK). The CGM was worn for the duration of the study period (10 days). Participants were given instructions to follow in the 24 hours ahead of each test day; avoid drinking alcohol, strenuous exercise and fast for 8 hr (no food or drink except water). On the morning of each test day participants were instructed to avoid smoking and use of tobacco products and consume a standardised amount of water. Baseline measures were conducted via a questionnaire booklet immediately before consumption of the test meal, and included subjective ratings of hunger, satiety, energy, mood and alertness. Participants were then instructed to consume test meals within a 15 min time window (0-15 min). Following consumption of the test meal, participants were asked to fast for 3 hr, avoid smoking or use of tobacco products, avoid strenuous exercise, avoid taking medications and were permitted to consume a standardised amount of water during this time. Further questionnaires were completed at 15, 60, 120 and 180 min. After completion of the 3 hr post-meal fast, participants reported the time they consumed their next meal and details of food consumed in their questionnaire booklet. When all four test days had been completed, participants returned their questionnaire booklets via prepaid return envelope. The postprandial sub-study was conducted between 6th and 17th May 2024.

### Test meals

A standardised high available carbohydrate breakfast was designed, consisting of white bread (128g; 57.6g carbohydrate (CHO), 3.2g fat, 11.5g protein, 2.9g fibre) and low fat spread (10-15g; 0.1g CHO, 3.5g fat, 0.1g protein, fibre not reported). The control test meal consisted of the standardised breakfast meal alone. The intervention test meal consisted of the standardised breakfast meal, in combination with the prebiotic blend (30g; 5.3g CHO, 7.7g fat, 5.5g protein, 9.0g fibre; ZOE ltd., UK). The nutrient composition of test meals is included in **Supplementary Table 8.**

### Continuous glucose monitoring

Interstitial glucose was measured every minute and aggregated into 15 minute readings, using Freestyle Libre Pro CGM (Abbott Diabetes Care, Alameda, CA, US). Glucose measurements were downloaded from the CGM onto the FreeStyle LibreLink mobile application (Abbott) by scanning the device with a smartphone containing the application download. Participants were provided with login details that linked their LibreLink application to the study practice account for retrieval of outcome data. Participants applied CGM devices 24hr before their first test meal, and data for the first 12 hr of CGM usage were discarded prior to analysis.

### Primary outcome

The primary outcome of the postprandial sub-study was the difference in peak postprandial glucose concentration (C-max) between the intervention and control test meals assessed using CGM-derived glucose concentration data.

### Secondary outcomes

Additional CGM derived metrics indicative of postprandial glycaemic response were analysed as secondary outcomes, including the difference in 2-h incremental area under the curve (2-h iAUC), time to max concentration (T-max), 2-3h dips below baseline (dips), and Time Course Analysis (i.e. Meal*Time interactions). Subjective ratings of satiety (hunger, fullness, desire to eat, satisfaction, prospective consumption), energy, mood (happiness, anxiety) and alertness were assessed using visual analogue scales (VAS; 0-100mm) ^52^. Time to next meal (min) and energy and macronutrient intake at next meal were assessed by a food diary included in the participant questionnaire booklet.

### Sample size calculations

For the chronic study, the primary outcome was based on the change in relative abundances of microbiome species previously identified for their associations with markers of cardiometabolic health^34^, from baseline to the 6-week endpoint. The study was powered to detect differences between groups in the primary outcome measure, using proprietary data collected within the ZOE commercial product. Based on a two-sided significance level (ɑ) of 0.05, with 85% power, a sample size of 102 participants per group (306 participants in total) was calculated. An anticipated attrition rate of 20-25% was applied based on rates in previous studies conducted by the research group, resulting in a total of 133 participants per group (399 participants in total).

The postprandial sub-study was powered to detect changes in the primary outcome (glucose C-max) using pilot data collected during the ZOE PREDICT 1 study (*unpublished data*)^58^. A within patient standard deviation of the difference in peak glucose concentration following a high carbohydrate vs a high fibre breakfast test meal was calculated (1.19 mmol/L). Based on a two-sided significance level (ɑ) of 0.05, with 80% power, a minimal detectable difference of 0.582 mmol/L, a sample size of 35 participants was calculated. Based on previous similar studies conducted by our research group, we anticipate a dropout rate of 15%, resulting in a total of 40 participants being required to take part in the crossover sub-study.

### Statistical analysis

Analysis was conducted using R Studio v2023.12.0 and Python v3.9.16 (package SciPy v1.11.4). Figures were created on Graphpad Prism Version 10.2.2. The statistical analysis plan was pre-registered on The Open Science Framework (https://osf.io/) prior to commencement of hypothesis testing.

Analysis of 6-week changes in primary and secondary outcomes were conducted on the ITT cohort (n = 349) and subgroups for selected metabolomic markers and gastrointestinal symptoms. For the primary outcome, we assessed gut microbiome composition using species-level taxonomic profiles of participants with samples available at baseline and 6-weeks (n = 321). The primary outcome measure was based on the specific bacterial species (n = 100) previously identified for their association with favourable (n = 50) or unfavourable (n = 50) cardiometabolic health markers^34^. Within each group, we identified species with a statistically significant difference in relative abundance values from baseline to 6-weeks using the Paired Wilcoxon signed-rank test. *P* values were corrected for multiple comparisons using the Benjamini–Hochberg false discovery rate (FDR). We then tested whether the significantly increasing species had significantly higher values of the “ZOE Microbiome Ranking 2024 (Cardiometabolic Health)” compared to the values of the significantly decreasing species within each group using the Mann-Whitney U-test (FDR corrected). Ranks are presented in box plots as median (IQR). We repeated the above analysis to determine the effect of the intervention on species previously identified for their association with favourable (n = 50) or unfavourable (n = 50) indices of diet quality “ZOE Microbiome Ranking 2024 (Diet)”^34^. As basic gut microbiome information we calculated alpha-diversity (observed richness, Shannon’s diversity index) and beta-diversity (unweighted-UniFrac) metrics. For alpha-diversity measures we assessed within group differences between baseline and 6-weeks using the paired Wilcoxon signed-rank test. For beta-diversity, differences between groups at baseline and 6-weeks were assessed using PERMANOVA.

Secondary outcome data was assessed for normality by visual inspection of histograms and the Shapiro-Wilk statistic. If outcome data was not normally distributed it was log_10_-transformed prior to analysis. To compare the effect of treatments on continuous outcomes (metabolites) a linear mixed effects model was applied, with participant ID as a random effect, and time and the interaction between time (within-subject factor) and treatment (between-subject factor) as fixed effects ^59^. Descriptive statistics are presented as mean, standard deviation (s.d.) for normally distributed variables, or geometric mean (95% confidence intervals, CI) for transformed variables. Changes in outcomes from baseline to 6-weeks are presented as mean (95% CI). For ordinal variables (gut symptoms, stool consistency), or data that could not be normalised by transformation (anthropometric measures, subjective emotions), change between baseline and 6-weeks was assessed using the Mann-Whitney test and presented as median (interquartile range, IQR). Differences between groups in categorical outcomes (stool frequency, sleep quality and quantity, and skin health) at 6-weeks were assessed using a chi-square test, and are presented as the number (n) and %.

For the postprandial sub-study, data was collected in duplicate, with the mean of the duplicate meals used for analysis. In instances where only one test meal data was available, the single meal response was included in the analysis. Summary statistics, including glucose C-Max, T-max, next meal data and iAUC for glucose and subjective outcomes, were analysed using a linear mixed effects model. The model included meal type and meal sequence as well as their interaction as fixed effects, while participant ID was incorporated as a random effect to account for individual variation. Normality of the model residuals was assessed visually using QQ plots and statistically tested using Shapiro-Wilk’s test. In cases where the residuals of the model were not normally distributed, even after data transformation, non-parametric tests were employed. A linear mixed-effects model was employed to analyse the time-course data, with fixed effects for timepoint, meal, and their interaction, and random intercepts to account for participant-specific effects. For all tests, the significance level was set at P < 0.05 (2-tailed).

## Declarations

### Ethics approval and consent to participate

Ethical approval was granted by King’s College London Research Ethics Committee (HR/DP-23/24-39673). All procedures were carried out in accordance with the Declaration of Helsinki (2013) and Good Clinical Practice. Informed consent was obtained from all volunteer participants.

### Funding

This research was funded by ZOE limited.

### Data availability

The study data can be released to bona fide researchers by submitting a research proposal approved by a subpanel of our scientific advisory board which meets once per month with independent members to assess proposals. The data may be pseudonymised or otherwise redacted to comply with the UK General Data Protection Regulation (GDPR). Researchers will be required to enter into a GDPR data sharing agreement before data may be released. Access request proposals should be sent to. The microbiome data will be made freely available via the EBI website (www.ebi.ac.uk)

### Authors’ contributions

J.W. and T.D.S. obtained the funding. F.Am., N.S., W.J.B., K.M.B., I.L., R.D., F.As., T.D.S., and S.E.B. contributed to the study design and developed the study concept. H.B., M.W., H.A.S., A.P., C.J., C.C., N.K. collected the data. A.C.C., H.B., K.M.B., and S.E.B. coordinated the study. M.W., A.A., H.A.S., A.P., J.C.P., E.P., A.R.L., C.J., F.G., F.As., and A.C.C. prepared and analysed the data. A.C.C., H.B., F.Am., A.A., H.A.S., W.J.B., F.As., T.D.S., and S.E.B. wrote the manuscript. All authors read and approved the final manuscript.

## Acknowledgements

We thank the participants of the BIOME study. We also thank G. Hadjigeorgiou, A. Majid, D. Boland, G. Cotrupi, L. Parker, M. Deldeniya, H.Warne, S. Gordon, M. Gallivan, R. DSouza and V. Zely for their assistance during the study and manuscript preparation. We thank Indi Supplements for their contribution toward the development of the prebiotic blend. This research was funded by ZOE Ltd; the study funder contributed, as part of the scientific advisory board, to study design, data collection and analysis, and the writing of the manuscript.

## Competing interests

T.D.S. and J.W. are co-founders of Zoe Ltd.. S.E.B., F.As., N.S. are consultants to ZOE Ltd., A.C.C., H.B., F.Am., M.W., A.A., H.A.S., A.P., W.J.B., K.M.B., J.C.P., A.R.L., C.J., C.C., N.K., I.L., F.G., and R.D., are or have been employees of ZOE Ltd. H.B., F.Am., A.A., M.W., H.A.S., A.P., W.J.B., K.M.B., J.C.P., A.R.L., I.L., F.G., F.As., J.W., N.S., S.E.B., and T.D.S. receive options with ZOE Ltd.

## Notes

### Clinical Trial

NCT06231706

### Author Declarations

Kings College London Research Ethics Committee (REC) of King's College London gave ethical approval for this work.

