## Supplementary Information for "A diverse high-fibre plant-based dietary intervention improves gut microbiome composition, gut symptoms, energy and hunger in healthy adults: a randomised controlled trial"

**Supplementary Table 1**. Nutrient intake in the BIOME study

| | Prebiotic blend | | | Probiotic | | | Control | | | *p-values* | |
|---|---|---|---|---|---|---|---|---|---|---|---|
| | *Baseline* (n=116) | *Endpoint* (n=107) | *Δ baseline-end* | *Baseline* (n=113) | *Endpoint* (n=107) | *Δ baseline-end* | *Baseline* (n=120) | *Endpoint* (n=110) | *Δ baseline-end* | Prebiotic blend vs control | Prebiotic blend vs probiotic |
| Energy (kcal/d) | 1738 ± 546 | 1770 ± 629 | 38 (-49, 125) | 1628 ± 442 | 1801 ± 551 | 161 (69, 252) | 1783 ± 527 | 1802 ± 609 | 12 (-88, 112) | 0.92 | 0.16 |
| Total carbohydrate (g/d) | 173 ± 75 | 176 ± 84 | 4 (-6, 14) | 170 ± 54 | 181 ± 59 | 9 (-1, 19) | 186 ± 70 | 188 ± 83 | 1 (-11, 14) | 0.94 | 0.77 |
| Total carbohydrate (%EI/d) | 40 ± 11 | 40 ± 10 | 0 (-1, 2) | 42 ± 9 | 41 ± 9 | -1 (-3, 0) | 42 ± 10 | 41 ± 10 | 0 (-2, 1) | 0.87 | 0.25 |
| Total sugar (g/d) | 65 ± 34 | 71 ± 47 | 5 (-1, 12) | 66 ± 28 | 67 ± 29 | 1 (-4, 6) | 75 ± 36 | 75 ± 44 | 0 (-7, 7) | 0.46 | 0.64 |
| Total sugar (%EI/d) | 15 ± 6 | 16 ± 7 | 1 (0, 2) | 16 ± 6 | 15 ± 5 | -1 (-2, 0) | 17 ± 6 | 17 ± 7 | 0 (-1, 1) | 0.41 | **0.04** |
| Total fat (g/d) | 76 ± 32 | 74 ± 31 | -3 (-8, 3) | 71 ± 25 | 73 ± 28 | 2 (-3, 7) | 76 ± 27 | 73 ± 30 | -4 (-9, 2) | 0.95 | 0.37 |
| Total fat (%EI/d) | 39 ± 9 | 37 ± 8 | -2 (-4, -1) | 39 ± 8 | 37 ± 8 | -2 (-4, -1) | 38 ± 8 | 36 ± 7 | -2 (-3, -1) | 0.96 | 0.99 |
| Saturated fat (g/d) | 27 ± 13 | 27 ± 13 | 0 (-3, 2) | 24 ± 10 | 26 ± 11 | 1 (-1, 3) | 27 ± 11 | 27 ± 13 | -1 (-3, 1) | 0.96 | 0.37 |
| Saturated fat (%EI/d) | 14 ± 4 | 14 ± 4 | -1 (-1, 0) | 13 ± 4 | 13 ± 3 | 0 (-1, 0) | 14 ± 4 | 13 ± 3 | -1 (-1, 0) | 0.94 | 0.97 |
| PUFA (g/d) | 13 ± 7 | 12 ± 6 | -1 (-2, 0) | 12 ± 5 | 12 ± 5 | 0 (-1, 1) | 13 ± 7 | 12 ± 5 | -2 (-3, 0) | 0.55 | 0.47 |
| PUFA (%EI/d) | 7 ± 3 | 6 ± 2 | -1 (-1, 0) | 7 ± 2 | 6 ± 2 | -1 (-1, 0) | 7 ± 3 | 6 ± 2 | -1 (-1, 0) | 0.99 | 0.86 |
| MUFA (g/d) | 29 ± 14 | 28 ± 13 | -1 (-3, 1) | 28 ± 11 | 29 ± 12 | 1 (-2, 3) | 29 ± 11 | 28 ± 13 | -1 (-3, 1) | 1 | 0.52 |
| MUFA (%EI/d) | 15 ± 5 | 14 ± 4 | -1 (-2, 0) | 15 ± 4 | 14 ± 4 | -1 (-2, 0) | 15 ± 4 | 14 ± 4 | -1 (-1, 0) | 0.75 | 0.99 |
| Protein (g/d) | 75 ± 26 | 72 ± 27 | -3 (-8, 2) | 71 ± 23 | 71 ± 21 | 0 (-4, 4) | 80 ± 32 | 73 ± 24 | -6 (-11, -1) | 0.56 | 0.49 |
| Protein (%EI/d) | 18 ± 4 | 17 ± 4 | -1 (-2, 0) | 18 ± 4 | 16 ± 4 | -1 (-2, -1 | 18 ± 5 | 17 ± 5 | -1 (-2, 0) | 1 | 0.72 |
| Fibre (g/d) | 17 ± 6 | 16 ± 7 | 0 (-2, 1) | 17 ± 6 | 17 ± 5 | -1 (-2, 0) | 18 ± 6 | 17 ± 7 | -2 (-3, 0) | 0.34 | 0.73 |
| Fibre (g/1000 kcal/d) | 10 ± 3 | 9 ± 4 | 0 (-1, 0) | 11 ± 3 | 10 ± 3 | -1 (-2, -1) | 11 ± 3 | 10 ± 3 | -1 (-2, 0) | 0.36 | 0.07 |
| Alcohol (g/d) | 15 ± 22 | 22 ± 28 | 9 (5, 12) | 10 ± 13 | 26 ± 33 | 15 (10, 20) | 11 ± 20 | 21 ± 24 | 9 (6, 12) | 0.97 | 0.07 |
| Alcohol (%EI/d) | 6 ± 8 | 9 ± 10 | 3 (2, 4) | 4 ± 5 | 9 ± 9 | 5 (4, 6) | 5 ± 8 | 8 ± 9 | 3 (2, 4) | 0.97 | 0.11 |

Data at baseline and endpoint are mean ± s.d., delta values are mean (95%CI). *P*-values are derived from an analysis of covariance (ANCOVA) with baseline values as a covariate. Values for fibre are derived from methods developed by the Association of Analytical Chemists (AOAC). Values at endpoint represent energy and macronutrient intake from background diet (not including intervention foods).

Kcal, kilocalories; %EI, percentage of total energy intake; MUFA, monounsaturated fatty acids; PUFA, polyunsaturated fatty acids; %EI, percentage of total energy intake

**Supplementary Table 2.** Secondary outcomes (categorical) in the BIOME study.

| | Prebiotic blend | | Probiotic | | Control | | *p-values* | |
|---|---|---|---|---|---|---|---|---|
| | *Baseline* (n = 116) | *Endpoint* (n = 107) | *Baseline* (n = 113) | *Endpoint* (n = 107) | *Baseline* (n = 120) | *Endpoint* (n = 110) | Prebiotic blend vs control | Prebiotic blend vs probiotic |
| **Stool frequency n (%)** | | | | | | | | |
| Once a week or less | 2 (1.7) | 0 (0) | 1 (0.0) | 1 (0.9) | 1 (0.8) | 1 (0.8) | | |
| Twice a week | 1 (0.9) | 3 (2.6) | 2 (1.8) | 1 (0.9) | 1 (0.8) | 3 (2.5) | | |
| Three or four times a week | 12 (10.3) | 4 (3.5)[a] | 13 (11.5) | 14 (12.4) | 24 (20) | 17 (14.2)[b] | | |
| Five or six times a week | 20 (17.2) | 9 (7.8) | 21 (18.6) | 15 (13.3) | 16 (13.3) | 17 (14.2) | | |
| Once a day | 58 (50) | 58 (50) | 52 (46) | 53 (46.9) | 54 (45) | 47 (39.2) | | |
| Twice a day | 20 (17.2) | 26 (22.4) | 21 (18.6) | 20 (17.7) | 19 (15.8) | 19 (15.8) | | |
| Three times a day | 2 (1.72) | 6 (5.2) | 3 (2.7) | 3 (2.7) | 5 (4.17) | 6 (5) | | |
| Four times a day | 1 (0.9) | 1 (0.9) | 0 (0) | 0 (0) | 0 (0) | 0 (0) | **0.040** | 0.098 |
| **Skin improvement n (%)** | | | | | | | | |
| Yes | | 2 (1.9) | | 0 (0) | | 1 (0.9) | | |
| No | | 11 (10.3) | | 8 (7.5) | | 11 (10) | | |
| Unsure | | 5 (4.7) | | 2 (1.9) | | 1 (0.9) | | |
| NA | | 89 (83.2) | | 97 (90.7) | | 97 (88.2) | 0.347 | 0.251 |
| **Sleep Quality n (%)**[+] | | | | | | | | |
| Very good | 7 (6.1) | 13 (12.1) | 10 (8.9) | 14 (13.1) | 18 (15) | 13 (11.8) | | |
| Fairly good | 65 (56.5) | 79 (73.8) | 71 (62.8) | 70 (65.4) | 63 (52.5) | 69 (62.7) | | |
| Fairly bad | 40 (34.8) | 13 (12.1) | 31 (27.4) | 23 (21.5) | 37 (30.8) | 27 (24.5) | | |
| Very bad | 3 (2.6) | 2 (1.9) | 1 (0.9) | 0 (0) | 2 (1.67) | 1 (0.9) | 0.127 | 0.155 |

| | | | | | | | | |
|---|---|---|---|---|---|---|---|---|
| **Sleep quantity n (%)**[+] | | | | | | | | |
| Less than 5hr | 1 (0.9) | 0 (0) | 1 (0.9) | 1 (0.9) | 4 (3.33) | 1 (0.9) | | |
| 5-6hr | 29 (25.2) | 18 (17) | 24 (21.2) | 19 (17.8) | 21 (17.5) | 17 (15.5) | | |
| 6-7hr | 44 (38.3) | 47 (44.3) | 41 (36.3) | 48 (44.9) | 44 (36.7) | 48 (43.6) | | |
| 7-8hr | 33 (28.7) | 32 (30.2) | 41 (36.3) | 31 (29) | 44 (36.7) | 37 (33.6) | | |
| 8-9hr | 8 (7.0) | 9 (8.5) | 6 (5.3) | 7 (6.5) | 6 (5) | 5 (4.6) | | |
| 9-10hr | 0 (0) | 0 (0) | 0 (0) | 0 (0) | 1 (0.83) | 1 (0.9) | | |
| 10-11hr | 0 (0) | 0 (0) | 0 (0) | 1 (0.94) | 0 (0) | 1 (0.9) | 0.613 | 0.806 |

Data are number, n, and percentage, %. *P*-values are a result of chi-square test; where comparisons between groups were significant, data within each row that do not share the same superscript letter are significantly different ($p<0.05$)

[+] Data for one participant is missing from the prebiotic group at baseline (n = 115)

**Supplementary Table 3.** Secondary outcomes (continuous) in the BIOME study.

| | **Prebiotic blend** | | | | | **Probiotic** | | | | | **Control** | | | | | *p-values* | |
|---|---|---|---|---|---|---|---|---|---|---|---|---|---|---|---|---|---|
| | n | *Baseline* | n | *Endpoint* | *Δ baseline-end* | n | *Baseline* | n | *Endpoint* | *Δ baseline-end* | n | *Baseline* | n | *Endpoint* | *Δ baseline-end* | Prebiotic blend vs control | Prebiotic blend vs probiotic |
| **Metabolomics**[a] | | | | | | | | | | | | | | | | | |
| ApoB (g/L)[+] | 106 | 0.91 (0.88, 0.95) | 97 | 0.89 (0.86, 0.92) | -0.01 (-0.03, 0.01) | 108 | 0.90 (0.87, 0.93) | 101 | 0.89 (0.85, 0.92) | 0.11 (-0.02, 0.02) | 106 | 0.88 (0.85, 0.92) | 100 | 0.87 (0.83, 0.91) | 0.00 (-0.03, 0.02) | 0.719 | 0.911 |
| GlycA (mmol/L)[+] | 103 | 0.80 (0.78, 0.81) | 95 | 0.79 (0.78, 0.81) | 0.00 (-0.02, 0.02) | 110 | 0.81 (0.79, 0.82) | 103 | 0.80 (0.79, 0.82) | 0.00 (-0.02, 0.01) | 107 | 0.79 (0.78, 0.80) | 100 | 0.80 (0.79, 0.82) | 0.02 (0.00, 0.03) | 0.513 | 0.526 |
| HbA1c (mmol/mol) | 100 | 33.88 (33.33, 34.43) | 93 | 33.55 (33.0, 34.11) | -0.04 (-0.44, 0.37) | 103 | 34.29 (33.78, 34.82) | 90 | 34.58 (34.02, 35.14) | 0.00 (-0.38, 0.38) | 103 | 33.89 (33.31, 34.47) | 96 | 34.01 (33.45, 34.57) | 0.00 (-0.34, 0.34) | 0.781 | 0.252 |
| Triglycerides (mmol/L)[+] | 105 | 1.17 (0.10, 1.24) | 96 | 1.09 (1.02, 1.07) | -0.04 (-0.13, 0.05) | 107 | 1.09 (1.02, 1.16) | 101 | 1.10 (1.03, 1.17) | 0.01 (-0.06, 0.08) | 107 | 1.11 (1.04, 1.19) | 101 | 1.14 (1.05, 1.23) | 0.06 (-0.03, 0.15) | 0.220 | 0.239 |
| Total cholesterol (mmol/L)[+] | 107 | 5.60 (5.44, 5.77) | 98 | 5.64 (5.50, 5.79) | 0.06 (-0.05, 0.17) | 109 | 5.62 (5.48, 5.78) | 102 | 5.62 (5.47, 5.78) | -0.01 (-0.13, 0.10) | 108 | 5.53 (5.37, 5.70) | 102 | 5.48 (5.31, 5.66) | -0.02 (-0.14, 0.10) | 0.350 | 0.660 |
| Omega-3 FAs[+] | 105 | 0.76 (0.72, 0.79) | 96 | 0.75 (0.71, 0.79) | 0.01 (-0.03, 0.04) | 110 | 0.75 (0.71, 0.78) | 103 | 0.76 (0.73, 0.80) | 0.02 (-0.01, 0.04) | 107 | 0.73 (0.70, 0.77) | 102 | 0.73 (0.69, 0.77) | 0.00 (-0.04, 0.04) | 0.693 | 0.612 |
| Total PUFA (mmol/L)[+] | 107 | 6.44 (6.29, 6.59) | 98 | 6.42 (6.27, 6.57) | -0.02 (-0.14, 0.10) | 110 | 6.33 (6.19, 6.47) | 103 | 6.36 (6.23, 6.48) | 0.02 (-0.07, 0.12) | 109 | 6.37 (6.21, 6.52) | 103 | 6.37 (6.20, 6.54) | 0.05 (-0.09, 0.18) | 0.834 | 0.676 |
| ApoA1 (g/L)[+] | 106 | 1.74 (1.69, 1.79) | 97 | 1.78 (1.73, 1.84) | 0.04 (0.00, 0.08) | 110 | 1.80 (1.75, 1.85) | 103 | 1.84 (1.79, 1.89) | 0.01 (-0.03, 0.05) | 110 | 1.78 (1.72, 1.83) | 104 | 1.80 (1.75, 1.86) | 0.01 (-0.02, 0.04) | 0.588 | 0.368 |
| Ratio of ApoB:ApoA1[+] | 106 | 0.52 (0.49, 0.54) | 97 | 0.49 (0.46, 0.52) | -0.01 (-0.03, 0.00) | 107 | 0.49 (0.47, 0.52) | 101 | 0.48 (0.46, 0.51) | 0.00 (-0.02, 0.01) | 108 | 0.49 (0.47, 0.52) | 102 | 0.48 (0.45, 0.51) | 0.00 (-0.02, 0.01) | 0.644 | 0.419 |
| Total cholesterol minus HDL-C (mmol/L) | 108 | 3.78 (3.63, 3.94) | 99 | 3.74 (3.60, 3.89) | -0.01 (-0.10, 0.09) | 109 | 3.74 (3.60, 3.88) | 102 | 3.71 (3.56, 3.86) | -0.01 (-0.10, 0.08) | 108 | 3.61 (3.44, 3.78) | 102 | 3.57 (3.39, 3.75) | 0.00 (-0.12, 0.12) | 0.390 | 0.898 |
| Remnant cholesterol (mmol/L)[+] | 108 | 1.64 (1.58, 1.71) | 99 | 1.62 (1.56, 1.69) | 0.00 (-0.04, 0.04) | 109 | 1.63 (1.57, 1.69) | 102 | 1.63 (1.57, 1.70) | 0.02 (-0.03, 0.06) | 107 | 1.57 (1.51, 1.64) | 101 | 1.56 (1.49, 1.63) | 0.01 (-0.04, 0.05) | 0.453 | 0.984 |
| VLDL-C (mmol/L)[+] | 106 | 0.62 (0.59, 0.65) | 97 | 0.59 (0.55, 0.62) | -0.02 (-0.04, 0.00) | 109 | 0.60 (0.57, 0.64) | 102 | 0.60 (0.56, 0.63) | 0.00 (-0.02, 0.02) | 105 | 0.59 (0.55, 0.62) | 99 | 0.57 (0.53, 0.61) | 0.00 (-0.03, 0.03) | 0.583 | 0.478 |
| Clinical LDL-C (mmol/L) | 108 | 3.02 (2.88, 3.16) | 99 | 2.97 (2.84, 3.10) | -0.01 (-0.09, 0.08) | 109 | 2.98 (2.86, 3.11) | 102 | 2.93 (2.80, 3.07) | -0.03 (-0.11, 0.05) | 105 | 2.92 (2.79, 3.06) | 99 | 2.79 (2.65, 2.95) | -0.09 (-0.18, 0.00) | 0.344 | 0.949 |
| LDL-C (mmol/L)[+] | 108 | 2.14 (2.05, 2.23) | 99 | 2.11 (2.03, 2.20) | 0.00 (-0.06, 0.05) | 109 | 2.11 (2.03, 2.19) | 102 | 2.07 (1.99, 2.16) | -0.03 (-0.08, 0.03) | 109 | 2.03 (1.91, 2.15) | 103 | 2.00 (1.89, 2.11) | -0.02 (-0.09, 0.05) | 0.380 | 0.699 |
| HDL-C (mmol/L) | 105 | 1.75 (1.69, 1.82) | 96 | 1.80 (1.73, 1.88) | 0.06 (0.01, 0.11) | 110 | 1.81 (1.74, 1.89) | 103 | 1.85 (1.78, 1.92) | 0.00 (-0.05, 0.05) | 110 | 1.81 (1.74, 1.88) | 104 | 1.81 (1.74, 1.88) | -0.01 (-0.05, 0.03) | 0.141 | 0.316 |

| | | | | | | | | | | | | | | | | | |
|---|---|---|---|---|---|---|---|---|---|---|---|---|---|---|---|---|---|
| Concentration of VLDL particles (mmol/L)+ | 106 | 0.00 (0.00, 0.00) | 97 | 0.00 (0.00, 0.00) | 0.00 (0.00, 0.00) | 110 | 0.00 (0.00, 0.00) | 103 | 0.00 (0.00, 0.00) | 0.00 (0.00, 0.00) | 108 | 0.00 (0.00, 0.00) | 102 | 0.00 (0.00, 0.00) | 0.00 (0.00, 0.00) | 0.285 | 0.233 |
| Concentration of LDL particles (mmol/L)+ | 106 | 0.00 (0.00, 0.00) | 97 | 0.00 (0.00, 0.00) | 0.00 (0.00, 0.00) | 110 | 0.00 (0.00, 0.00) | 103 | 0.00 (0.00, 0.00) | 0.00 (0.00, 0.00) | 108 | 0.00 (0.00, 0.00) | 102 | 0.00 (0.00, 0.00) | 0.00 (0.00, 0.00) | 0.547 | 0.925 |
| Cholesterol in very large HDL (mmol/L) | 106 | 0.10 (0.08, 0.12) | 97 | 0.11 (0.09, 0.12) | 0.01 (0.00, 0.01) | 109 | 0.11 (0.10, 0.12) | 102 | 0.12 (0.11, 0.13) | 0.00 (-0.01, 0.01) | 110 | 0.12 (0.11, 0.13) | 104 | 0.11 (0.10, 0.12) | 0.00 (-0.01, 0.00) | 0.272 | 0.236 |
| Concentration of HDL particles (mmol/L) | 107 | 0.02 (0.02, 0.02) | 98 | 0.02 (0.02, 0.02) | 0.00 (0.00, 0.00) | 110 | 0.02 (0.02, 0.02) | 103 | 0.02 (0.02, 0.02) | 0.00 (0.00, 0.00) | 109 | 0.02 (0.02, 0.02) | 104 | 0.02 (0.02, 0.02) | 0.00 (0.00, 0.00) | 0.341 | 0.25 |
| Cholesterol in large HDL (mmol/L) | 106 | 0.38 (0.29, 0.50) | 97 | 0.39 (0.30, 0.52) | 0.03 (0.01, 0.06) | 110 | 0.46 (0.40, 0.53) | 103 | 0.51 (0.46, 0.56) | 0.00 (-0.03, 0.03) | 109 | 0.50 (0.46, 0.55) | 104 | 0.48 (0.43, 0.53) | -0.01 (-0.03, 0.01) | 0.060 | 0.263 |
| Cholesterol in medium HDL (mmol/L) | 105 | 0.65 (0.63, 0.67) | 96 | 0.68 (0.65, 0.71) | 0.03 (0.01, 0.05) | 110 | 0.66 (0.63, 0.69) | 103 | 0.68 (0.66, 0.71) | 0.00 (-0.02, 0.02) | 110 | 0.66 (0.63, 0.68) | 104 | 0.66 (0.64, 0.69) | 0.00 (-0.01, 0.02) | 0.112 | 0.211 |
| Cholesterol in small HDL (mmol/L) | 108 | 0.49 (0.48, 0.50) | 99 | 0.49 (0.48, 0.50) | 0.00 (-0.01, 0.01) | 110 | 0.49 (0.49, 0.50) | 103 | 0.49 (0.49, 0.50) | 0.00 (-0.01, 0.01) | 108 | 0.48 (0.47, 0.49) | 102 | 0.48 (0.47, 0.49) | 0.00 (-0.01, 0.01) | 0.681 | 0.508 |
| Cholesterol in very large VLDL (mmol/L)+ | 107 | 0.03 (0.03, 0.04) | 98 | 0.02 (0.02, 0.03) | 0.00 (-0.01, 0.00) | 110 | 0.03 (0.02, 0.03) | 103 | 0.03 (0.02, 0.03) | 0.00 (0.00, 0.00) | 110 | 0.03 (0.02, 0.03) | 104 | 0.02 (0.02, 0.03) | 0.00 (0.00, 0.01) | 0.304 | 0.349 |
| Triglycerides in very large VLDL (mmol/L)+ | 107 | 0.06 (0.04, 0.08) | 98 | 0.04 (0.03, 0.06) | 0.00 (-0.02, 0.01) | 110 | 0.03 (0.02, 0.05) | 103 | 0.03 (0.02, 0.05) | 0.00 (-0.01, 0.01) | 110 | 0.04 (0.02, 0.05) | 104 | 0.03 (0.02, 0.05) | 0.02 (0.00, 0.04) | 0.380 | 0.369 |
| Cholesterol in large VLDL (mmol/L)+ | 106 | 0.06 (0.05, 0.07) | 97 | 0.05 (0.04, 0.06) | -0.01 (-0.01, 0.00) | 110 | 0.06 (0.05, 0.07) | 103 | 0.05 (0.05, 0.06) | 0.00 (-0.01, 0.01) | 110 | 0.05 (0.05, 0.06) | 104 | 0.05 (0.04, 0.06) | 0.01 (0.00, 0.02) | 0.455 | 0.225 |
| Triglycerides in large VLDL (mmol/L)+ | 107 | 0.15 (0.14, 0.17) | 98 | 0.13 (0.12, 0.15) | -0.01 (-0.02, 0.01) | 110 | 0.14 (0.12, 0.15) | 103 | 0.12 (0.11, 0.14) | -0.01 (-0.02, 0.01) | 109 | 0.14 (0.12, 0.16) | 103 | 0.12 (0.10, 0.16) | 0.01 (-0.01, 0.03) | 0.740 | 0.572 |
| Cholesterol in medium VLDL (mmol/L) | 108 | 0.17 (0.16, 0.18) | 99 | 0.16 (0.15, 0.17) | 0.00 (-0.01, 0.00) | 109 | 0.16 (0.15, 0.17) | 102 | 0.16 (0.15, 0.17) | 0.00 (-0.01, 0.00) | 107 | 0.15 (0.14, 0.16) | 101 | 0.14 (0.13, 0.16) | 0.00 (-0.01, 0.00) | 0.485 | 0.854 |
| Triglycerides in medium VLDL (mmol/L)+ | 106 | 0.27 (0.25, 0.29) | 97 | 0.24 (0.23, 0.27) | -0.01 (-0.03, 0.01) | 110 | 0.25 (0.23, 0.27) | 103 | 0.24 (0.22, 0.26) | -0.01 (-0.03, 0.01) | 110 | 0.26 (0.24, 0.28) | 104 | 0.25 (0.23, 0.28) | 0.02 (-0.01, 0.05) | 0.495 | 0.652 |
| Cholesterol in small VLDL (mmol/L) | 108 | 0.14 (0.13, 0.15) | 99 | 0.13 (0.12, 0.14) | 0.00 (-0.01, 0.00) | 109 | 0.13 (0.13, 0.14) | 102 | 0.13 (0.12, 0.14) | 0.00 (0.00, 0.01) | 106 | 0.12 (0.11, 0.13) | 100 | 0.12 (0.11, 0.13) | 0.00 (0.00, 0.01) | 0.252 | 0.374 |
| Triglycerides in small VLDL (mmol/L)+ | 106 | 0.14 (0.14, 0.15) | 97 | 0.13 (0.13, 0.14) | -0.01 (-0.02, 0.00) | 110 | 0.14 (0.13, 0.15) | 103 | 0.13 (0.13, 0.15) | 0.00 (-0.01, 0.01) | 109 | 0.14 (0.13, 0.15) | 103 | 0.14 (0.13, 0.15) | 0.01 (0.00, 0.02) | 0.106 | 0.344 |
| Cholesterol in very small VLDL (mmol/L) | 108 | 0.18 (0.17, 0.19) | 99 | 0.18 (0.17, 0.19) | 0.00 (-0.01, 0.00) | 109 | 0.18 (0.18, 0.19) | 102 | 0.19 (0.18, 0.19) | 0.01 (0.00, 0.01) | 106 | 0.18 (0.17, 0.18) | 100 | 0.17 (0.17, 0.18) | 0.00 (0.00, 0.00) | 0.594 | 0.359 |
| Triglycerides in very small VLDL (mmol/L)+ | 107 | 0.07 (0.06, 0.07) | 98 | 0.06 (0.06, 0.06) | 0.00 (-0.01, 0.00) | 110 | 0.07 (0.06, 0.07) | 103 | 0.07 (0.07, 0.07) | 0.00 (0.00, 0.01) | 106 | 0.06 (0.06, 0.07) | 100 | 0.07 (0.06, 0.07) | 0.00 (0.00, 0.01) | **0.006** | **0.016** |
| Cholesterol in IDL (mmol/L) | 107 | 0.98 (0.94, 1.02) | 98 | 0.99 (0.,95, 1.03) | 0.02 (-0.01, 0.05) | 109 | 1.00 (0.97, 1.04) | 102 | 1.02 (0.98, 1.06) | 0.01 (-0.01, 0.04) | 108 | 0.97 (0.94, 1.01) | 102 | 0.97 (0.93, 1.01) | 0.00 (-0.03, 0.03) | 0.586 | 0.789 |
| Triglycerides in IDL (mmol/L)+ | 107 | 0.11 (0.10, 0.11) | 98 | 0.10 (0.10, 0.11) | 0.00 (0.00, 0.00) | 110 | 0.11 (0.10, 0.11) | 103 | 0.11 (0.11, 0.11) | 0.01 (0.00, 0.01) | 105 | 0.10 (0.10, 0.10) | 99 | 0.10 (0.10, 0.11) | 0.01 (0.00, 0.01) | **0.009** | **0.016** |

| | | | | | | | | | | | | | | | | | |
|---|---|---|---|---|---|---|---|---|---|---|---|---|---|---|---|---|---|
| Cholesterol in large LDL (mmol/L) | 106 | 1.41 (1.36, 1.48) | 97 | 1.41 (1.36, 1.47) | 0.01 (-0.03, 0.05) | 110 | 1.41 (1.36, 1.46) | 103 | 1.39 (1.33, 1.45) | -0.01 (-0.05, 0.02) | 109 | 1.37 (1.32, 1.44) | 104 | 1.33 (1.26, 1.40) | -0.03 (-0.07, 0.01) | 0.383 | 0.853 |
| Cholesterol in medium LDL (mmol/L)+ | 107 | 0.50 (0.47, 0.53) | 98 | 0.48 (0.46, 0.51) | -0.01 (-0.02, 0.01) | 110 | 0.49 (0.47, 0.52) | 103 | 0.48 (0.46, 0.51) | -0.01 (-0.02, 0.01) | 110 | 0.46 (0.41, 0.51) | 104 | 0.47 (0.43, 0.50) | 0.00 (-0.02, 0.02) | 0.438 | 0.990 |
| Cholesterol in small LDL (mmol/L)+ | 108 | 0.21 (0.20, 0.22) | 99 | 0.21 (0.20, 0.21) | 0.00 (-0.01, 0.00) | 110 | 0.21 (0.20, 0.21) | 103 | 0.20 (0.19, 0.21) | 0.00 (-0.01, 0.00) | 110 | 0.20 (0.19, 0.21) | 104 | 0.20 (0.19, 0.21) | 0.00 (-0.01, 0.01) | 0.563 | 0.905 |
| **Symptom severity (individual)[b]** | | | | | | | | | | | | | | | | | |
| Hunger pains | 116 | 1.40 (0.70) | 107 | 1.22 (0.48) | -0.18 (-0.30, -0.06) | 113 | 1.41 (0.66) | 107 | 1.21 (0.45) | -0.18 (-0.27, -0.09) | 120 | 1.50 (0.90) | 110 | 1.32 (0.77) | -0.15 (-0.27, -0.04) | 0.694 | 0.841 |
| Nausea | 116 | 1.11 (0.43) | 107 | 1.13 (0.41) | 0.02 (-0.04, 0.08) | 113 | 1.19 (0.71) | 107 | 1.07 (0.25) | -0.13 (-0.25, 0.01) | 120 | 1.22 (0.65) | 110 | 1.17 (0.57) | -0.03 (-0.16, 0.10) | 0.570 | 0.219 |
| Heartburn | 116 | 1.45 (1.00) | 107 | 1.14 (0.44) | -0.29 (-0.42, -0.16) | 113 | 1.21 (0.57) | 107 | 1.06 (0.27) | -0.12 (-0.21, -0.03) | 120 | 1.25 (0.66) | 110 | 1.21 (0.62) | -0.05 (-0.16, -0.05) | **0.045** | 0.146 |
| Acid reflux | 116 | 1.28 (0.90) | 107 | 1.18 (0.45) | -0.09 (-0.20, 0.01) | 113 | 1.2 (0.68) | 107 | 1.09 (0.32) | -0.04 (-0.10, 0.03) | 120 | 1.10 (0.40) | 110 | 1.15 (0.51) | 0.05 (-0.03, 0.12) | 0.306 | 0.971 |
| Rumbling stomach | 116 | 1.49 (0.76) | 107 | 1.23 (0.45) | -0.26 (-0.39, -0.14) | 113 | 1.67 (0.94) | 107 | 1.34 (0.58) | -0.33 (-0.47, -0.19) | 120 | 1.57 (0.84) | 110 | 1.47 (0.80) | -0.06 (-0.19, 0.07) | 0.060 | 0.600 |
| Loose stool | 116 | 1.34 (0.66) | 107 | 1.23 (0.54) | -0.12 (-0.24, -0.01) | 113 | 1.43 (0.88) | 107 | 1.2 (0.48) | -0.22 (-0.36, -0.09) | 120 | 1.37 (0.66) | 110 | 1.36 (0.75) | 0.00 (-0.15, 0.15) | 0.383 | 0.573 |
| Hard stool | 116 | 1.30 (0.58) | 107 | 1.20 (0.52) | -0.11 (-0.21, -0.02) | 113 | 1.31 (0.60) | 107 | 1.29 (0.64) | 0 (-0.10, 0.10) | 120 | 1.34 (0.65) | 110 | 1.33 (0.78) | -0.01 (-0.15, 0.13) | 0.625 | 0.241 |
| Constipation | 116 | 1.37 (0.67) | 107 | 1.13 (0.44) | -0.26 (-0.37, -0.16) | 113 | 1.31 (0.6) | 107 | 1.34 (0.75) | 0.04 (-0.09, -0.17) | 120 | 1.35 (0.67) | 110 | 1.35 (0.77) | 0.02 (-0.10, 0.14) | **0.019** | **0.007** |
| Diarrhoea | 116 | 1.27 (0.68) | 107 | 1.17 (0.56) | -0.10 (-0.21, 0.00) | 113 | 1.25 (0.84) | 107 | 1.12 (0.47) | -0.10 (-0.22, 0.02) | 120 | 1.23 (0.63) | 110 | 1.22 (0.67) | -0.02 (-0.16, 0.12) | 0.520 | 0.735 |
| Urgent movement | 116 | 1.53 (0.98) | 107 | 1.40 (0.76) | -0.14 (-0.26, -0.02) | 113 | 1.36 (0.85) | 107 | 1.21 (0.51) | -0.12 (-0.26, 0.01) | 120 | 1.48 (0.95) | 110 | 1.35 (0.89) | -0.14 (-0.27, 0.0) | 0.727 | 0.681 |
| Bowel emptying | 116 | 1.47 (0.77) | 107 | 1.36 (0.59) | -0.14 (-0.27, -0.01) | 113 | 1.56 (0.82) | 107 | 1.41 (0.63) | -0.13 (-0.27, -0.01) | 120 | 1.62 (1.09) | 110 | 1.57 (0.94) | 0.03 (-0.09, 0.15) | 0.226 | 0.874 |
| Bloated | 116 | 1.81 (1.06) | 107 | 1.58 (0.95) | -0.23 (-0.39, -0.08) | 113 | 1.87 (1.03) | 107 | 1.46 (0.73) | -0.38 (-0.52, -0.24) | 120 | 1.83 (1.16) | 110 | 1.76 (1.16) | -0.03 (-0.20, 0.14) | 0.367 | 0.133 |
| Burping | 116 | 1.44 (0.89) | 107 | 1.27 (0.61) | -0.18 (-0.29, -0.06) | 113 | 1.35 (0.85) | 107 | 1.21 (0.50) | -0.12 (-0.26, -0.01) | 120 | 1.28 (0.65) | 110 | 1.24 (0.51) | -0.02 (-0.10, 0.06) | 0.164 | 0.573 |
| Flatulence | 116 | 1.98 (1.00) | 107 | 1.68 (0.73) | -0.33 (-0.49, -0.17) | 113 | 2.08 (1.14) | 107 | 1.60 (0.78) | -0.49 (-0.66, -0.31) | 120 | 1.77 (0.99) | 110 | 1.68 (0.90) | -0.05 (-0.66, -0.31) | **0.048** | 0.267 |
| Abdominal pain | 116 | 1.41 (0.75) | 107 | 1.29 (0.67) | -0.11 (-0.24, 0.02) | 113 | 1.62 (1.09) | 107 | 1.21 (0.50) | -0.39 (-0.55, -0.24) | 120 | 1.49 (0.91) | 110 | 1.45 (0.88) | -0.01 (-0.17, 0.15) | 0.635 | 0.068 |
| **Waist circumference[c] (cm)** | | | | | | | | | | | | | | | | | |

| | | | | | | | | | | | | | | | | |
|---|---|---|---|---|---|---|---|---|---|---|---|---|---|---|---|---|
| Male | 29 | 97.0 (16.0) | 27 | 94.0 (18.5) | 0.00 (5.0) | 27 | 98.0 (16.5) | 23 | 92.0 (13.5) | -1.00 (10.5) | 30 | 97.5 (15.7) | 28 | 93.0 (15.5) | -2.00 (16.5) | 0.336 | 0.297 |
| Female | 86 | 85.5 (18.7) | 79 | 85.0 (16.0) | -1.00 (6.0) | 86 | 84.0 (17.8) | 84 | 85.0 (17.0) | -0.50 (4.0) | 90 | 85.0 (18.7) | 92 | 87.0 (18.0) | 1.00 (5.75) | 0.092 | 0.907 |
| **Weight[c] (kg)** | 115 | 73.0 (22.7) | 106 | 73.2 (21.9) | 0.20 (1.89) | 113 | 74.5 (22.0) | 107 | 73.5 (21.9) | 0.00 (1.84) | 120 | 75.0 (17.1) | 110 | 75.3 (19.7) | 0.25 (1.95) | 0.495 | **0.018** |
| **Stool consistency[c] (BSFS)** | 116 | 4 (1) | 107 | 4 (1) | 0 (1) | 113 | 4 (1) | 107 | 4 (1) | 0 (1) | 120 | 4 (1) | 110 | 4 (1) | 0 (1.75) | 0.334 | 0.943 |

[a] Data are geometric mean (95%CI) for baseline and endpoint values, and mean (95%CI) for delta values; *p*-values are the result of a linear mixed model, with time and treatment*time as fixed effects, and a random intercept for participant ID.

[b] Data were not normally distributed, but are presented as mean (SD), and mean (95%CI) for delta values to illustrate precise changes between the groups for each outcome (as median values did not indicate the direction); *p*-values are a result of a Mann-Whitney U-test on median change from baseline values.

[c] Data are median (IQR); *p*-values are a result of a Mann-Whitney U-test on median change from baseline values.

[+] *p*-values are the result of statistical tests on log-transformed data

ApoB, apolipoprotein B; GlycA, glycoprotein acetyls; HbA1c, glycated haemoglobin; FAs, fatty acids, PUFA, polyunsaturated fatty acids; ApoA1, apolipoprotein A1; BSFS, bristol stool form scale

**Supplementary Table 4.** Subgroup analysis of selected secondary outcomes in the BIOME study

| | **Prebiotic blend** | | | | | **Probiotic** | | | | | **Control** | | | | | ***p-values*** | |
|---|---|---|---|---|---|---|---|---|---|---|---|---|---|---|---|---|---|
| | n | *Baseline* | n | *Endpoint* | *Δ baseline-end* | n | *Baseline* | n | *Endpoint* | *Δ baseline-end* | n | *Baseline* | n | *Endpoint* | *Δ baseline-end* | Prebiotic blend vs control | Prebiotic blend vs probiotic |
| **Metabolomics[a]** | | | | | | | | | | | | | | | | | |
| ApoB (g/L) | 39 | 1.14 (1.11, 1.17) | 33 | 1.07 (1.03, 1.12) | -0.06 (-0.09, -0.03) | 34 | 1.17 (1.13, 1.22) | 30 | 1.13 (1.09, 1.18) | -0.02 (-0.06, 0.02) | 34 | 1.13 (1.10, 1.17) | 29 | 1.14 (1.09, 1.19) | 0.02 (-0.04, 0.07) | 0.068 | 0.136 |
| GlycA (mmol/L) | 34 | 0.90 (0.88, 0.92) | 29 | 0.85 (0.82, 0.89) | -0.04 (-0.08, -0.01) | 38 | 0.92 (0.90, 0.94) | 34 | 0.88 (0.85, 0.91) | -0.04 (-0.06, -0.01) | 35 | 0.90 (0.88, 0.92) | 33 | 0.90 (0.87, 0.94) | 0.01 (-0.02, 0.04) | 0.075 | 0.384 |
| LDL-C (mmol/L) | 39 | 2.79 (2.69, 2.89) | 32 | 2.57 (2.45, 2.69) | -0.22 (-0.31, -0.13) | 34 | 2.83 (2.73, 2.94) | 30 | 2.65 (2.52, 2.79) | -0.15 (-0.25, -0.04) | 36 | 2.76 (2.66, 2.86) | 31 | 2.69 (2.54, 2.84) | -0.04 (-0.18, 0.09) | 0.229 | 0.553 |
| Triglycerides (mmol/L) | 37 | 1.74 (1.61, 1.88) | 32 | 1.53 (1.35, 1.72) | -0.14 (-0.36, 0.07) | 32 | 1.80 (1.66, 1.95) | 30 | 1.51 (1.36, 1.68) | -0.2 (-0.37, -0.03) | 37 | 1.79 (1.66, 1.93) | 33 | 1.71 (1.51, 1.95) | 0.03 (-0.19, 0.26) | 0.403 | 0.904 |
| **Symptom severity[b] (domains)** | | | | | | | | | | | | | | | | | |
| Acid reflux | 17 | 3.15 (1.28) | 15 | 1.73 (0.70) | -1.37 (-1.87, -0.86) | 13 | 2.46 (0.78) | 10 | 1.30 (0.48, -0.90) | -0.9 (-1.31, -0.49) | 10 | 2.50 (0.67) | 10 | 1.90 (1.47) | -0.50 (-1.16, 0.16) | 0.091 | 0.377 |
| Abdominal pain | 14 | 2.12 (0.36) | 13 | 1.62 (0.71) | -0.51 (-0.76, -0.26) | 21 | 2.33 (0.61) | 20 | 1.40 (-.41, -0.87) | -0.87 (-1.13, -0.61) | 17 | 2.59 (0.95) | 14 | 1.74 (0.84) | -0.81 (-1.07, -0.55) | 0.283 | 0.206 |
| Indigestion | 41 | 2.37 (0.50) | 39 | 1.71 (0.61) | -0.67 (-0.82, -0.51) | 34 | 2.61 (0.66) | 31 | 1.73 (0.55, -0.88) | -0.88 (-1.07, -0.69) | 31 | 2.56 (0.75) | 28 | 2.1 (0.89) | -0.36 (-0.55, -0.16) | 0.119 | 0.154 |
| Diarrhoea | 22 | 2.42 (0.58) | 21 | 1.81 (1.81) | -0.63 (-0.95, -0.32) | 12 | 3.11 (1.48) | 11 | 1.70 (0.96, -1.27) | -1.27 (-2.24, -0.31) | 18 | 2.43 (0.66) | 16 | 1.92 (1.06) | -0.56 (-1.02, -0.11) | 0.865 | 0.390 |
| Constipation | 22 | 2.26 (0.42) | 21 | 1.56 (0.63) | -0.71 (-0.97, -0.46) | 20 | 2.27 (0.41) | 18 | 1.70 (0.95, -0.50) | -0.50 (-0.81, -0.19) | 22 | 2.61 (0.67) | 19 | 1.96 (1.19) | -0.54 (-0.90, -0.18) | 0.816 | 0.440 |
| **Symptom severity[b] (individual)** | | | | | | | | | | | | | | | | | |
| Hunger pains | 34 | 2.35 (0.60) | 31 | 1.39 (0.56) | -1.00 (-1.24, -0.76) | 37 | 2.24 (0.55) | 34 | 1.44 (0.61) | -0.76 (-0.93, -0.60) | 39 | 2.54 (0.94) | 34 | 1.71 (1.17) | -0.82 (-1.07, -0.58) | 0.301 | 0.144 |
| Rumbling stomach | 42 | 2.36 (0.66) | 38 | 1.39 (0.55) | -1.00 (-1.21, -0.79) | 52 | 2.46 (0.87) | 49 | 1.53 (0.71) | -0.92 (-1.14, -0.70) | 49 | 2.39 (0.76) | 44 | 1.82 (1.04) | -0.52 (-0.78, -0.27) | 0.008 | 0.355 |
| Bowel emptying | 39 | 2.41 (0.68) | 37 | 1.68 (0.71) | -0.76 (-1.02, -0.49) | 44 | 2.43 (0.70) | 41 | 1.63 (0.70) | -0.78 (-0.98, -0.58) | 45 | 2.64 (1.23) | 39 | 2.08 (1.24) | -0.46 (-0.67, -0.25) | 0.168 | 0.970 |
| Bloating | 54 | 2.74 (0.89) | 50 | 2.00 (1.20) | -0.74 (-1.01, -0.47) | 59 | 2.66 (0.84) | 54 | 1.76 (0.85) | -0.91 (-1.11, -0.71) | 54 | 2.83 (1.06) | 50 | 2.26 (1.35) | -0.48 (-0.77, -0.19) | 0.357 | 0.366 |

| | | | | | | | | | | | | | | | | |
|---|---|---|---|---|---|---|---|---|---|---|---|---|---|---|---|---|
| Gas | 72 | 2.58 (0.82) | 67 | 1.88 (0.75) | -0.73 (-0.93, -0.53) | 71 | 2.72 (0.97) | 67 | 1.79 (0.86) | -0.94 (-1.16, -0.72) | 60 | 2.55 (0.87) | 55 | 2.04 (1.02) | -0.44 (-0.62, -0.25) | 0.105 | 0.175 |
| Abdominal pain | 33 | 2.45 (0.67) | 29 | 1.59 (0.82) | -0.90 (-1.14, -0.65) | 38 | 2.84 (1.13) | 37 | 1.51 (0.69) | -1.24 (-1.16, -0.91) | 39 | 2.51 (1.00) | 36 | 1.69 (0.86) | -0.72 (-0.95, -0.49) | 0.485 | 0.263 |
| Loose stool | 31 | 2.29 (0.64) | 29 | 1.48 (0.74) | -0.83 (-1.11, -0.55) | 33 | 2.48 (1.03) | 31 | 1.42 (0.72) | -1.03 (-1.38, -0.68) | 34 | 2.29 (0.58) | 31 | 1.45 (0.86) | -0.84 (-1.10, -0.58) | 0.805 | 0.656 |
| Hard stool | 29 | 2.21 (0.49) | 27 | 1.56 (0.85) | -0.67 (-0.95, -0.38) | 28 | 2.25 (0.52) | 26 | 1.58 (1.03) | -0.62 (-0.88, -0.35) | 31 | 2.32 (0.60) | 29 | 1.55 (1.09) | -0.72 (-1.05, -0.40) | 0.422 | 0.881 |
| Constipation | 32 | 2.34 (0.55) | 31 | 1.35 (0.71) | -1.00 (-1.24, -0.76) | 28 | 2.25 (0.52) | 26 | 1.62 (1.10) | -0.62 (-0.97, -0.26) | 32 | 2.31 (0.64) | 29 | 1.79 (1.05) | -0.45 (-0.78, -0.11) | 0.033 | 0.114 |
| Urgent movement | 38 | 2.63 (1.08) | 34 | 1.88 (1.07) | -0.82 (-1.07, -0.58) | 27 | 2.52 (1.12) | 24 | 1.50 (0.78) | -0.96 (-1.43, -0.49) | 35 | 2.63 (1.11) | 32 | 1.72 (1.33) | -0.97 (-1.22, -0.72) | 0.216 | 0.699 |

[a] Subgroup analysis was performed within the top tertile concentrations (using baseline values) of key measures of lipids and inflammation. Data are geometric mean (95%CI) for baseline and endpoint values, and mean (95%CI) for delta values; *p*-values are the result of a linear mixed model, with time and treatment*time as fixed effects, and a random intercept for participant ID. Data was log-transformed prior to statistical analysis.

[b] Subgroup analysis was performed on participants who reported symptoms at baseline with a severity score ≥2 (individual symptoms and domains). Only individual symptoms that were reported by ≥25% of participants at baseline were assessed to ensure adequate sample size (n = 10 symptoms). Data were not normally distributed, but are presented as mean (SD), and mean (95%CI) for delta values to illustrate precise changes between the groups for each outcome (as median values did not indicate the direction); *p*-values are a result of a Mann-Whitney U-test on median change from baseline values.

ApoB, apolipoprotein B; GlycA, glycoprotein acetyls

**Supplementary Table 5.** Postprandial interstitial glucose responses and time to next meal in the BIOME postprandial crossover sub-study.

| | High CHO test meal | High CHO test meal + prebiotic blend | *p*-value |
|---|---|---|---|
| Glucose C-max (mmol/L)[a] | 7.28 (6.65, 8.28) | 7.50 (6.71, 8.54) | 0.495 |
| Glucose T-max (mins) | 63 (19) | 62 (23) | 0.639 |
| Glucose 2h iAUC (mmol/l x 2h) | 140.1 (96.4) | 130.5 (94.0) | 0.165 |
| Glucose 2-3h dip (%) | -2.06 (12.6) | -2.22 (13.9) | 0.275 |
| Time to next meal (mins) | 75 (94) | 87 (70) | 0.116 |

Data are mean (s.d.) unless otherwise stated. *p*-value from linear mixed effects model analysis of log transformed data (fixed effects for meal, meal sequence and their interaction, and a random intercept for subject ID); [a]Data are median (IQR), *p*-value from non-parametric test (paired Wilcoxon sign-rank test). n = 34 participants included in analyses (crossover design).

C-max, maximum glucose concentration; T-max. Time to maximum glucose concentration; iAUC, incremental area under the curve

**Supplementary Table 6**. Prebiotic blend (Daily30+) ingredients

| **Prebiotic blend ingredients** |
|---|
| Onion, garlic, red beetroot, carrot, baobab fruit, buckthorn, thyme, parsley, rosemary, white mushroom, lions mane, reishu, chaga, shiitake, cordyceps, maitke, tremella, almonds, walnuts, hazelnuts, chicory root inulin, nutritional yeast, flax seed, sunflower seed, chia seed, pumpkin seed, hemp seed, grape seed, tumeric, cumin, red lentil flakes puffed quinoa |

**Supplementary Table 7**. Nutrient composition of study treatments in the BIOME study

| | **Prebiotic blend (30g)** | **Control (28g)** |
|---|---|---|
| Energy (kcal) | 126.9 | 125.7 |
| Fat (g) | 7.7 | 3.8 |
| of which saturates (g) | 0.8 | 0.4 |
| Carbohydrate (g) | 5.3 | 19.3 |
| of which sugars (g) | 1.0 | 1.6 |
| Fibre (g) | 9.0 | 1.3 |
| Protein (g) | 5.5 | 3.0 |
| Salt (g) | 0.0 | 0.3 |

Kcal, kilocalories; g, grams

**Supplementary Table 8**. Nutrient composition of test meals in the postprandial sub-study

| | **High carbohydrate breakfast (control)** | **High carbohydrate breakfast + prebiotic blend** |
|---|---|---|
| Energy (kcal) | 342.5 | 469.4 |
| Fat (g) | 6.7 | 14.4 |
| of which saturates (g) | 1.4 | 2.1 |
| Carbohydrate (g) | 57.7 | 63.0 |
| of which sugars (g) | 2.9 | 3.8 |
| Fibre (g) | 2.9 | 11.9 |
| Protein (g) | 11.6 | 17.1 |
| Salt (g) | 1.4 | 1.4 |

Kcal, kilocalories; g, grams

**Supplementary Figure 1**. Overview of microbiome diversity measures

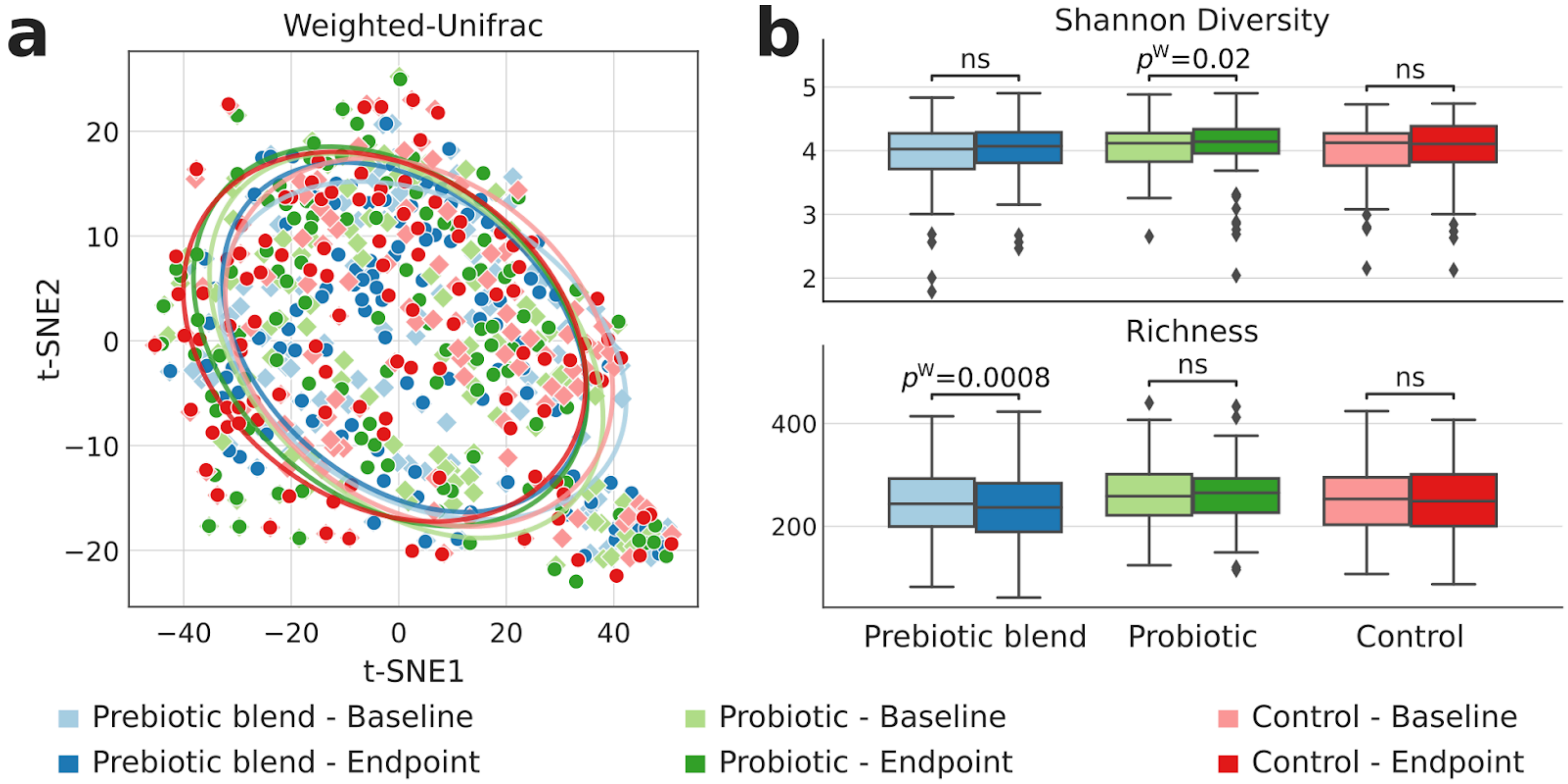


**a** Beta-diversity calculated with the weighted-UniFrac measure, shows no separation between the baseline microbiome composition across groups (PERMANOVA *p*-value = 0.584)., while endpoint microbiome compositions show significant differences (PERMANOVA *p*-value = 0.020). PERMANOVA analysis performed within each group comparing baseline with endpoint microbiome composition, showed significant differences only for the prebiotic blend (prebiotic blend *p*-value = 0.0297, probiotic *p*-value = 0.3267, control *p*-value = 0.0594). **b** Alpha diversity calculated both with the Shannon's diversity index (top) and richness (bottom), showed that Shannon's diversity index tended to increase across all three groups (significant only in probiotic group, Wilcoxon's *p*-value = 0.0203), while richness showed a slightly lower number of detected species at endpoint across all three groups (significant only in the prebiotic blend group, Wilcoxon's *p*-value = 0.0008).

**Supplementary Figure 2.** Time-course analysis of postprandial glucose metrics.

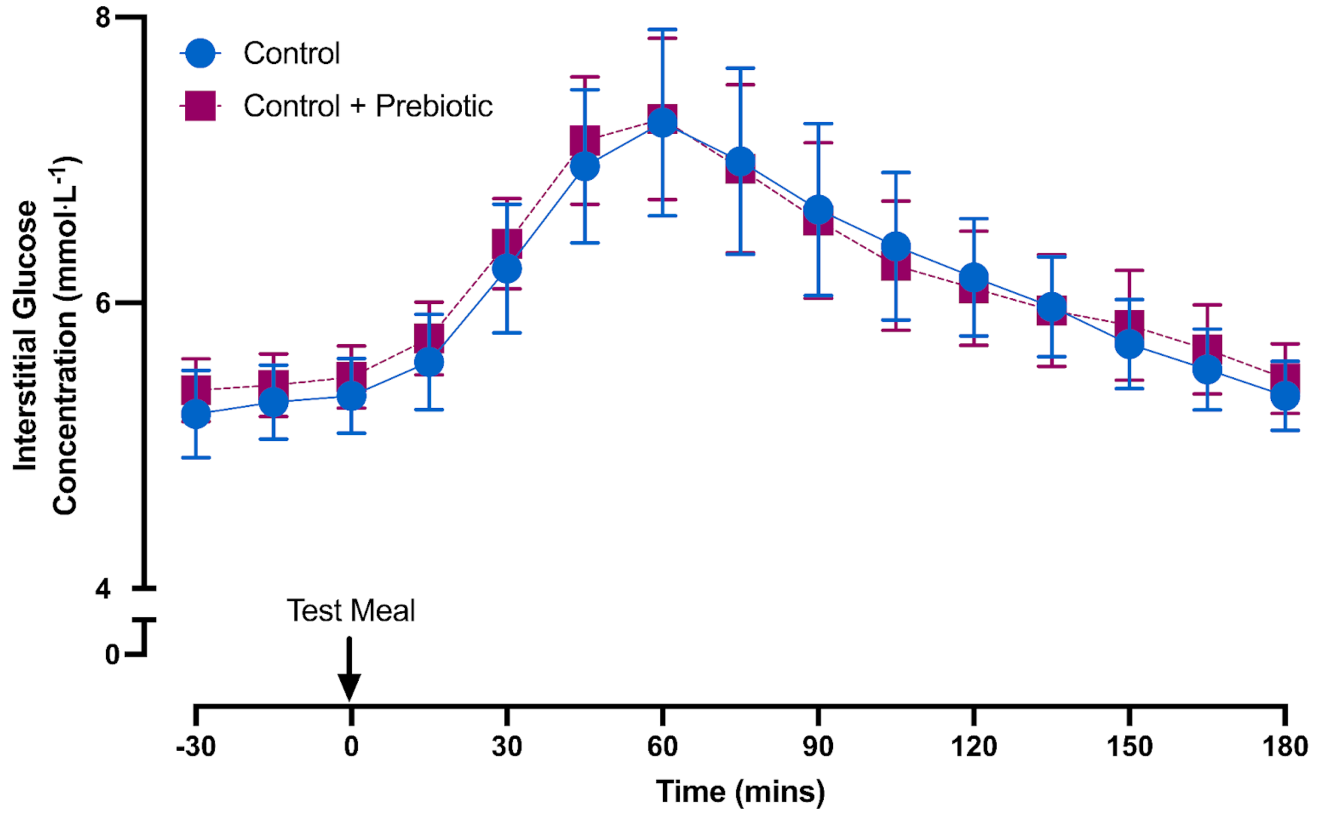


Time course of postprandial interstitial glucose concentration (3h) following a standardized high carbohydrate meal alone (blue) and in combination with a prebiotic blend (pink). No significant difference was observed between the test meals (linear mixed-effects model, with fixed effects for timepoint, meal, and their interaction, and a random intercept for subject ID). n = 34 participants (crossover design).

**Supplementary Figure 3**. Time-course analysis of postprandial subjective outcomes

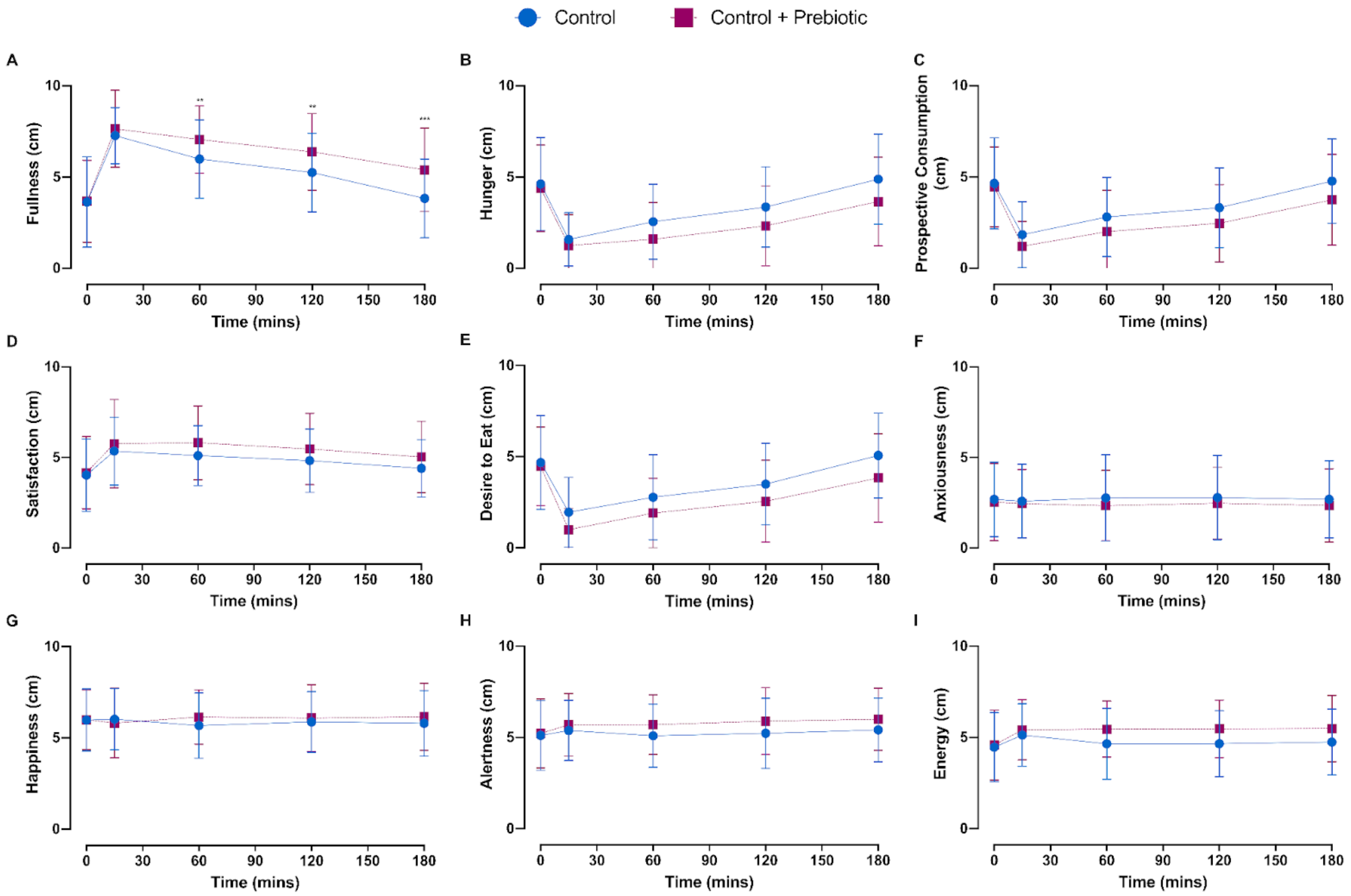


Time course of postprandial subjective feelings (3h) following a standardized high carbohydrate meal alone (blue) and in combination with a prebiotic blend (pink). There was a significant meal*time interaction for fullness ($p = 0.020$). No other significant interactions were observed between the test meals (linear mixed-effects model, with fixed effects for timepoint, meal, and their interaction, and a random intercept for subject ID). n = 34 participants included in analyses (crossover design).
